# Early patient-level quality of care and in-hospital outcomes among preterm and low-birth-weight newborns in Western Kenya: a multicentre retrospective record-review study

**DOI:** 10.64898/2026.11.16.26368257

**Authors:** Morris Senghor Shisanya, Lucy Natecho Namusonge, Catherine Mutonyi Simiyu, Tecla Cecilia Kirwa, Pauline Lusaria Waswa

## Abstract

Preterm and low-birth-weight newborns account for most neonatal deaths in sub-Saharan Africa, and survival depends on the quality and timeliness of care delivered after admission as well as on access to a facility. Quality is usually measured at facility level, and patient-level measures that combine early and later care processes are vulnerable to bias because newborns must survive in order to receive later care. We assessed whether better documented early care was associated with lower in-hospital mortality among preterm and low-birth-weight newborns in eight facilities in Western Kenya. In this multicentre retrospective record review, 291 source newborn records yielded 287 unique records, of which 275 met eligibility criteria and 268 had a calculable early-care patient-level quality score. This study-defined score covered thermal protection, skilled attendance and clinical review, assessment and monitoring, and medicines and fluids, each item scored as not done, partially done or fully done, with not-applicable items excluded from denominators. In-hospital survival status was known for 224 newborns, of whom 57 (25.4%) died before discharge; newborns referred out or still admitted were not counted as survivors. Modified Poisson regression with robust variance estimated risk ratios. Documented early care was high (median 93.5%, interquartile range 83.3–100), while kangaroo mother care was the weakest domain (median 87.5%). Early-care quality was not associated with in-hospital mortality after adjustment for gestational age, multiple pregnancy, birth location, facility and admission severity (adjusted risk ratio 1.03 per 10 percentage points, 95% confidence interval 0.82–1.29), and this null result persisted across nine sensitivity analyses. In contrast, continuing-care quality appeared strongly protective (0.81, 0.74–0.89), a pattern consistent with newborns surviving longer accruing more documented care rather than with a benefit of care itself.

## Introduction

Preterm birth is the leading cause of death in children under five years of age. An estimated 13.4 million babies were born preterm in 2020, and preterm and small newborns account for the majority of the world’s 2.3 million annual neonatal deaths, with sub-Saharan Africa and southern Asia bearing most of that burden [1,2]. Kenya’s neonatal mortality rate stands at 21 deaths per 1,000 live births, and complications of prematurity remain a leading contributor [3].

Survival for these newborns depends on more than reaching a health facility. The interventions that determine outcome — thermal protection from the moment of birth, skilled assessment, respiratory support, structured monitoring, and appropriate fluids and antimicrobials — must be delivered correctly and within a narrow time window after admission [4,5]. Where they are delayed or omitted, admission to a facility confers less benefit than its availability implies. This is the reasoning behind the shift in global newborn policy from measuring access to measuring the quality of care actually received [6,7].

Most quality measurement in low- and middle-income settings nevertheless describes facilities rather than newborns. Health facility assessments quantify whether equipment, medicines, guidelines and staff are present, and such assessments have been conducted at scale across East Africa [8,9]. Structural readiness is necessary but not sufficient: a functional warmer does not establish that a given newborn was kept warm, and a facility scoring highly on oxygen availability may still fail to monitor saturation in an individual newborn. Patient-level process measures, derived from what was documented in each newborn’s own record, sit closer to the care that newborn actually received, and are the level at which Donabedian’s process dimension of quality was originally defined [10].

Kenya has an unusually strong platform for such measurement. The KEMRI-coordinated Clinical Information Network links structured newborn admission records across hospitals to audit and feedback, and has generated large multisite analyses of newborn care and outcomes [11,12,13]. Yet published patient-level analyses have concentrated on individual processes — admission temperature, oxygen use, antibiotic prescribing — rather than on composite measures spanning the domains of early newborn care, and the relationship between documented composite process quality and in-hospital mortality among preterm and low-birth-weight newborns has been examined less often [14].

There is a methodological reason for caution in such analyses that has received insufficient attention. A composite quality score computed over an entire admission necessarily includes processes that can only occur if the newborn survives long enough to receive them: daily weight monitoring, sustained kangaroo mother care, feeding progression, discharge counselling and a discharge summary. A newborn who dies six hours after admission cannot accrue these items and will score poorly, not because care was deficient but because the opportunity to deliver and document it did not arise. Any association between such a score and survival is therefore partly structural. This is the same class of problem as immortal time bias in pharmacoepidemiology, in which exposure classification depends on surviving a period during which the outcome could occur [15,16]. Restricting the exposure to care processes that should occur during initial assessment and stabilisation is a direct, prespecified way of reducing it.

We therefore conducted a multicentre retrospective review of newborn clinical records from health facilities in Western Kenya to determine whether higher documented early patient-level quality of care was associated with lower in-hospital mortality among preterm and low-birth-weight newborns. We hypothesised that higher early-care quality would be associated with lower mortality. Secondary objectives were to describe patient-level quality across eight care domains, to examine variation across facilities and newborn risk groups, and to examine relationships between documented quality and complications, length of stay, weight change and feeding at discharge. Because the contrast between an early-care score and a whole-admission score is itself informative about the bias described above, both were analysed and compared.

## Materials and methods

### Study design and setting

This was a multicentre retrospective review of individual newborn clinical records, conducted in health facilities in Bungoma County, Western Kenya. Bungoma County operates a tiered public health system in which health centres and sub-county hospitals provide maternity services and stabilise and refer newborns requiring more than basic care, while the county referral hospital provides inpatient newborn care and receives referrals from across the county. Newborn records were abstracted from nine facilities; eligible preterm or low-birth-weight newborns were admitted at eight of them: one level 5 county referral hospital, six level 4 hospitals and one level 3 health centre. Reporting follows the STROBE statement for observational studies [17]; the completed checklist is provided as S1 Checklist.

### Data source and study population

Data were abstracted between 6 May and 26 June 2026 from newborn clinical records, admission registers, monitoring charts, treatment sheets and discharge or death summaries, using a structured instrument administered on tablets by trained research assistants. The instrument recorded maternal and obstetric history, newborn characteristics at birth and admission, condition at admission, care processes during admission, complications, and outcome. Records related to newborns born between July 2024 and May 2026. All admissions of preterm or low-birth-weight newborns identified in the source registers during the abstraction period were reviewed; no sampling within facilities was applied.

### Eligibility criteria

Newborns were eligible if they met at least one of the following: recorded gestational age below 37 completed weeks; a recorded preterm classification; birth weight below 2,500 g; or a recorded low-birth-weight classification. Records were excluded if they were confirmed duplicates not selected as the primary record, or if they contained neither usable gestational-age or preterm information nor usable birth-weight or low-birth-weight information, so that eligibility could not be determined. Records were not excluded solely because covariates were incomplete, provided the primary exposure and outcome were valid.

### Data management and quality assurance

Before analysis the dataset underwent independent cleaning, deduplication and validation. Each raw worksheet was reconciled cell-by-cell against the original electronic export; facility names, which were recorded as free text, were standardised using a crosswalk mapping every observed alias to a standard facility name and level; potential duplicates were identified using facility, record identifier, dates and newborn characteristics and classified as confirmed duplicates, documented multiple births or distinct records. Within confirmed duplicate groups the record with the highest weighted completeness was retained and singular non-conflicting values were recovered from complementary records; contradictory values were never merged. Documented multiple births were retained as separate newborns.

### Patient-level quality-of-care assessment

Care processes were assessed using a study-defined patient-level quality score (PLQS) based on prespecified clinical-care indicators drawn from national and World Health Organization guidance for the care of small and sick newborns. The score comprises 31 indicators grouped into eight domains: thermal protection; skilled attendance and clinical review; assessment and monitoring; medicines and fluids; feeding and nutrition; kangaroo mother care; infection prevention and control; and referral, escalation and continuity of care. Each indicator was scored 0 (not done or not documented), 1 (partially done, delayed or inconsistently documented) or 2 (fully and appropriately done and documented), with an explicit not-applicable option for indicators that did not apply to a given newborn.

Domain and composite scores were calculated as the sum of observed applicable item scores divided by the maximum achievable score for those applicable items, multiplied by 100. Not-applicable responses were excluded from denominators and were never recoded to zero. No PLQS item score was statistically completed at any stage. The PLQS is a study-defined measure constructed for this study; it has not undergone formal external psychometric validation and is not described as validated.

### Early-care PLQS: the primary exposure

The primary exposure was the early-care PLQS, calculated over the four domains comprising care processes that should reasonably occur during initial assessment and stabilisation: thermal protection, skilled attendance and clinical review, assessment and monitoring, and medicines and fluids. Items dependent on surviving long enough to receive later care were excluded by design, specifically daily weight and feeding monitoring, sustained kangaroo mother care documentation, discharge planning, discharge summaries and feeding at discharge. We verified that no discharge-dependent or transition-dependent indicator contributed to the primary exposure. The early-care PLQS was analysed as a continuous score from 0 to 100, modelled per 10-percentage-point increment, and secondarily in categories defined by the observed distribution.

For comparison, a continuing-care PLQS was calculated over the remaining four domains, and an overall PLQS over all applicable items. Both were analysed descriptively and as secondary exposures, and the contrast between them and the early-care score was used to assess the magnitude of opportunity-to-receive-care bias.

### Outcome definitions

The primary outcome was in-hospital death, defined as death before discharge (coded 1) compared with discharge alive (coded 0). Newborns referred out of the facility, still admitted at the time of abstraction, or with no recorded disposition were not classified as survivors and were excluded from the primary mortality analysis, because their final survival status could not be verified.

Secondary outcomes were: any complication documented during admission; sepsis during admission; hypothermia documented at admission; discharge alive; length of stay in days; weight change in grams between the baseline weight and discharge weight; average daily weight gain; human-milk feeding at discharge, defined as exclusive breastfeeding or expressed breast milk against other feeding modes; and referral out of the facility. All outcome definitions were taken from the audited dataset and no clinical outcome was inferred from absence of documentation.

### Covariates

Prespecified covariates were selected conceptually rather than by automated procedures. Core perinatal covariates were gestational age in completed weeks, multiple pregnancy, birth location (inborn against outborn) and facility. Newborn sex was not recorded in the abstraction instrument and could not be included; this is acknowledged as a limitation. Admission severity covariates were 5-minute Apgar score, hypothermia at admission, suspected sepsis at admission and respiratory distress at admission. All severity covariates referred to condition at presentation rather than to complications arising later in the admission, so that no post-exposure variable entered the models.

Gestational age and birth-weight category are strongly correlated markers of maturity. Gestational age was used in the primary adjustment set; birth-weight category replaced it in a prespecified sensitivity model, and the two were not entered together.

### Analysis populations

Four populations were prespecified. The quality-of-care cohort comprised eligible newborns with a calculable early-care PLQS. The mortality cohort comprised those with a definitively known in-hospital survival status. The discharged-survivor cohort comprised newborns discharged alive, and was used for length of stay, weight change, average daily weight gain and feeding at discharge. The 24-hour survivor cohort excluded newborns who died within 24 hours of admission where valid admission and outcome times permitted this determination, and was used for sensitivity analysis.

### Statistical analysis

Continuous variables are summarised as mean with standard deviation where approximately symmetric and as median with interquartile range where skewed; categorical variables as counts and percentages. Baseline comparisons between newborns who died and those discharged alive are presented with standardised mean differences rather than significance tests, since the purpose was to describe imbalance rather than to test it.

The primary analysis used modified Poisson regression with robust (sandwich) variance to estimate risk ratios and 95% confidence intervals for in-hospital death [18]. Three prespecified models were fitted: Model 1, unadjusted; Model 2, adjusted for gestational age, multiple pregnancy, birth location and facility; and Model 3, additionally adjusted for 5-minute Apgar score, hypothermia at admission and suspected sepsis at admission. Before fitting, the ratio of mortality events to candidate parameters was assessed and the facility term examined for sparse strata.

With eight facilities and mortality events concentrated in three of them, facility fixed effects at the individual-facility level produced complete separation for the group of facilities recording no deaths. Following the prespecified hierarchy, facility was therefore modelled as a binary term contrasting the two referral hospitals with operational newborn units against all other facilities, which eliminated sparse strata while retaining the conceptual distinction. Respiratory distress at admission was documented in 121 of 224 newborns in the mortality cohort and preceded 56 of 57 deaths, producing near-complete separation; including it yielded an unstable estimate and it was therefore excluded from the primary Model 3, with its crude and adjusted estimates reported separately.

Secondary binary outcomes were analysed by modified Poisson regression adjusted for gestational age, multiple pregnancy, birth location, facility and 5-minute Apgar score. Length of stay was highly overdispersed and was analysed by Poisson regression with robust variance, yielding an adjusted ratio of means. Weight change and average daily weight gain were analysed by linear regression with robust standard errors among discharged survivors with valid paired weights and biologically plausible values. Secondary and domain-level analyses are exploratory; the primary hypothesis concerns early-care PLQS and in-hospital mortality alone, and no multiplicity adjustment was applied to the prespecified primary model.

Nonlinearity of the exposure was examined by adding a quadratic term and, separately, a restricted cubic spline with three knots at the 10th, 50th and 90th centiles. Analyses were conducted in Python 3 using pandas, NumPy and SciPy.

### Missing data

Missingness in outcomes and PLQS item scores was never statistically completed. Covariates not recorded in the source records were completed once during the independent data audit using regression-based completion models; in the mortality cohort these were maternal age (49 of 224 records), antenatal visits (69), multiple pregnancy (13), 5-minute Apgar score (25), hypothermia at admission (14), hypoglycaemia at admission (24), respiratory distress at admission (8) and suspected sepsis at admission (8). The primary analysis used these completed covariate values. Gestational age, which the audit did not complete, was missing in 3 of 224 records (1.3%); it was imputed 20 times by chained equations, drawing regression parameters from their approximate posterior distribution and including the exposure, all model covariates and the outcome, and estimates were pooled using Rubin’s rules [19]. A complete-case analysis for gestational age is reported alongside. Two further analyses examined the influence of covariate completion: a complete-case analysis restricted to newborns with every Model 3 covariate recorded in the source records, and pooling across the 20 completed covariate datasets supplied with the audited data package. Source-record missingness for every analytical variable is given in S1 Table.

### Sensitivity and bias-control analyses

Five prespecified bias-control analyses were conducted. First, the early-care PLQS rather than the overall PLQS served as the primary mortality exposure. Second, we confirmed that no discharge-dependent or transition item contributed to that exposure. Third, the primary model was repeated among 24-hour survivors. Fourth, the analysis was restricted to newborns with at least three of the four early-care domains contributing valid information. Fifth, inborn and outborn newborns were analysed separately. Additional sensitivity analyses substituted birth-weight category for gestational age, and repeated the primary model using the overall and continuing-care scores as exposures. Interaction between the early-care PLQS and early preterm status (below 32 weeks), and between the early-care PLQS and referred-in status, was tested using multiplicative interaction terms.

### Ethics statement

Ethical approval was granted by the Masinde Muliro University of Science and Technology Institutional Scientific and Ethics Review Committee (MMUST-ISERC), approval number MMUST/ISERC/023/2026, correspondence reference MMU/COR: 40312 Vol 6(01), dated 2 February 2026 and valid from 2 February 2026 to 2 February 2027, for the study ’Health Systems Readiness and Quality of Care for Preterm Births in Western Kenya’. A national research licence was issued by the National Commission for Science, Technology and Innovation (NACOSTI), Kenya: licence number NACOSTI/P/26/4184987, reference 168774, issued 17 February 2026 and valid until 17 February 2027, for research in Bungoma, Kenya. County research authorisation was granted by the County Government of Bungoma, Ministry of Health, Office of the County Director of Health, reference CG/BGM/CDH/RESRC/VOL.1, dated 24 February 2026.

Access to routine clinical records was undertaken in accordance with the approved protocol and the requirements of MMUST-ISERC and the participating health authorities. Records were accessed within the facilities by trained research assistants. No names, telephone numbers or addresses of mothers or newborns were abstracted. The abstracted dataset contained facility newborn file numbers and dates and times of birth, admission and discharge or death, to which the authors had access during data cleaning; these were used only for duplicate resolution and the derivation of time intervals, and were removed from the analytical and shared datasets. Data were stored on password-protected devices accessible only to the study team, results are reported in aggregate, and the record-level dataset shared with this article is de-identified (see Data availability). MMUST-ISERC waived the requirement for individual informed consent for this retrospective review of routinely collected records; abstraction was conducted with the permission of the facility in-charges.

## Results

### Cohort derivation

The source dataset contained 291 newborn records. Four confirmed duplicates were excluded, leaving 287 unique records. Twelve newborns met neither the preterm nor the low-birth-weight criterion and were excluded, leaving 275 eligible newborns. A further seven had insufficient applicable information to calculate an early-care PLQS, giving a quality-of-care cohort of 268 newborns from eight facilities (Fig 1).

**Fig 1.**
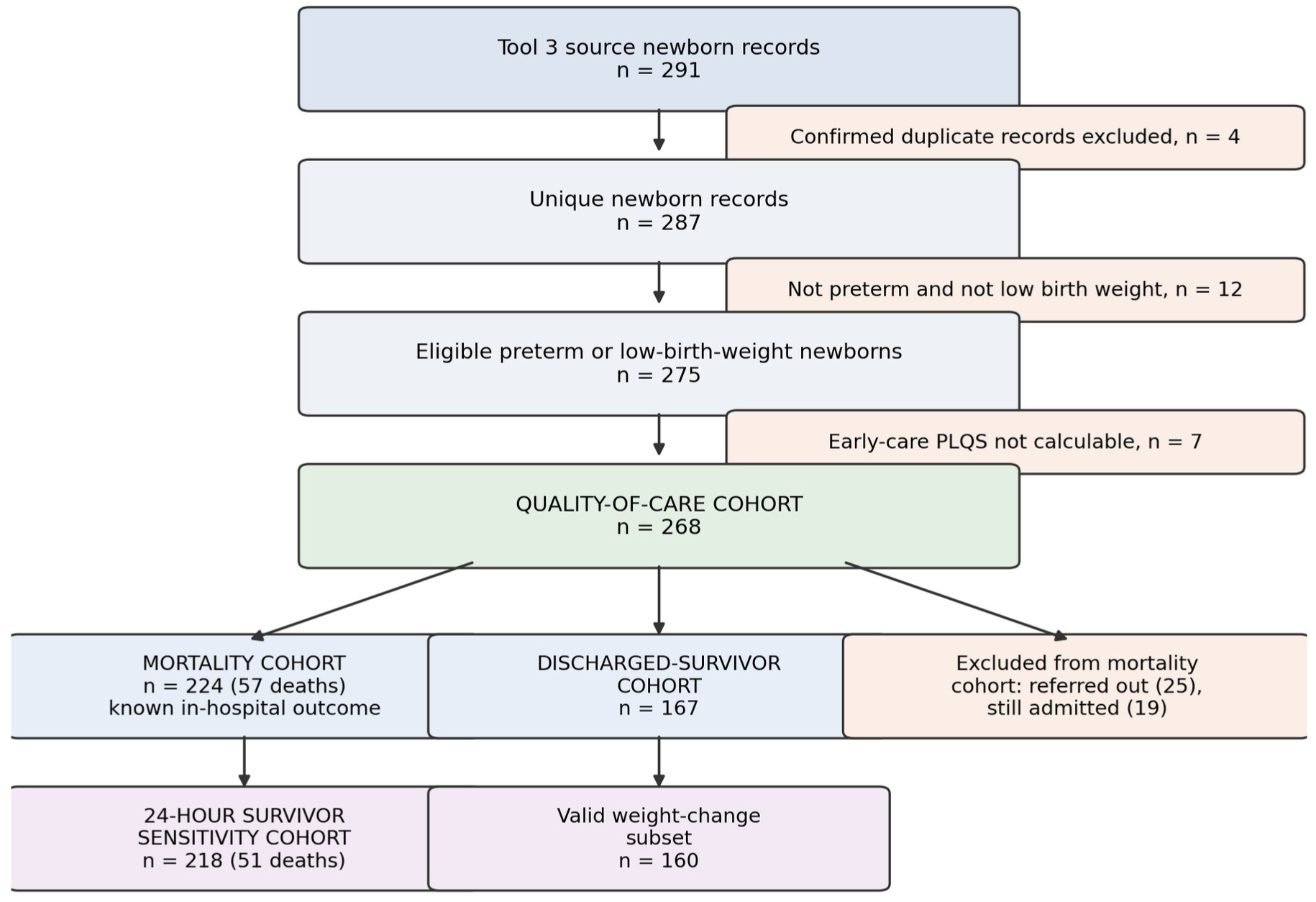
Cohort derivation. Flow of newborn records from the source dataset through duplicate removal, eligibility assessment and exclusions to the quality-of-care cohort, and thence to the mortality, discharged-survivor and 24-hour survivor sensitivity cohorts. PLQS, patient-level quality score

Within this cohort, in-hospital survival status was definitively known for 224 newborns, who formed the mortality cohort. The remaining 44 were excluded from the primary mortality analysis: 25 were referred out of the facility and 19 were still admitted at the time of abstraction, and none could be verified as having survived to discharge. The discharged-survivor cohort comprised 167 newborns, and the 24-hour survivor sensitivity cohort 218 newborns after excluding six who died within 24 hours of admission.

### Baseline maternal, obstetric and newborn characteristics

Of the 224 newborns in the mortality cohort, 57 (25.4%) died before discharge. Newborns who died differed markedly from those discharged alive on maturity and admission condition, and minimally on maternal characteristics (Table 1). Median gestational age was 28 weeks (IQR 27–32) among newborns who died against 34 weeks (IQR 32–35) among those discharged alive (standardised mean difference 1.34), and median birth weight was 1,200 g (IQR 950–1,500) against 1,900 g (IQR 1,600–2,100) (standardised difference 1.80). Extremely preterm birth accounted for 17 of 57 deaths (29.8%) but only 1 of 167 survivors (0.6%).

**Table 1.** Maternal, obstetric and newborn characteristics of the mortality cohort overall and by in-hospital survival status.

| Characteristic | Total (n=224) | Discharged alive (n=167) | Died (n=57) | SMD | Missing n |
|---|---|---|---|---|---|
| **Maternal and obstetric** |  |  |  |  |  |
| Maternal age (years) | 25 (22–32) | 25 (22–32) | 24 (20–34) | 0.00 | 49 |
| Antenatal visits | 1 (1–1) | 1 (1–1) | 1 (0–1) | 0.96 | 69 |
| Four or more antenatal visits | 18 (8.0) | 18 (10.8) | 0 (0.0) | 0.49 | 69 |
| Multiple pregnancy | 67 (29.9) | 47 (28.1) | 20 (35.1) | -0.15 | 13 |
| Caesarean delivery | 44 (19.6) | 36 (21.6) | 8 (14.0) | 0.20 | 0 |
| Maternal hypertensive disorder | 30 (13.4) | 21 (12.6) | 9 (15.8) | -0.09 | 26 |
| Maternal infection recorded | 27 (12.1) | 19 (11.4) | 8 (14.0) | -0.08 | 0 |
| Maternal anaemia | 5 (2.2) | 3 (1.8) | 2 (3.5) | -0.11 | 30 |
| Antenatal corticosteroids received | 20 (8.9) | 17 (10.2) | 3 (5.3) | 0.18 | 90 |
| **Newborn characteristics** |  |  |  |  |  |
| Gestational age (weeks) | 33 (30–35) | 34 (32–35) | 28 (27–32) | 1.34 | 3 |
| Prematurity category |  |  |  |  | 3 |
| Extremely preterm (<28 weeks) | 18 (8.0) | 1 (0.6) | 17 (29.8) | -0.89 |  |
| Very preterm (28–31 weeks) | 54 (24.1) | 30 (18.0) | 24 (42.1) | -0.55 |  |
| Moderate to late preterm (32–36 weeks) | 148 (66.1) | 134 (80.2) | 14 (24.6) | 1.34 |  |
| Term (≥37 weeks) | 1 (0.4) | 0 (0.0) | 1 (1.8) | -0.19 |  |
| Birth weight (g) | 1800 (1430–2025) | 1900 (1600–2100) | 1200 (950–1500) | 1.80 | 1 |
| Birth-weight category |  |  |  |  | 1 |
| Extremely low (<1000 g) | 19 (8.5) | 1 (0.6) | 18 (31.6) | -0.93 |  |
| Very low (1000–1499 g) | 41 (18.3) | 17 (10.2) | 24 (42.1) | -0.78 |  |
| Low (1500–2499 g) | 153 (68.3) | 138 (82.6) | 15 (26.3) | 1.37 |  |
| ≥2500 g | 10 (4.5) | 10 (6.0) | 0 (0.0) | 0.36 |  |
| 5-minute Apgar score | 8 (7–9) | 8 (7–9) | 7 (5–8) | 0.76 | 25 |
| Inborn | 164 (73.2) | 127 (76.0) | 37 (64.9) | 0.25 | 0 |
| Referred in | 51 (22.8) | 32 (19.2) | 19 (33.3) | -0.33 | 0 |
| **Condition at admission** |  |  |  |  |  |
| Hypothermia | 71 (31.7) | 35 (21.0) | 36 (63.2) | -0.95 | 14 |
| Hypoglycaemia | 41 (18.3) | 22 (13.2) | 19 (33.3) | -0.49 | 24 |
| Respiratory distress | 121 (54.0) | 65 (38.9) | 56 (98.2) | -1.66 | 8 |
| Suspected sepsis | 44 (19.6) | 25 (15.0) | 19 (33.3) | -0.44 | 8 |
| Jaundice | 24 (10.7) | 19 (11.4) | 5 (8.8) | 0.09 | 11 |
| **Facility** |  |  |  |  |  |
| Bumula Sub-County Hospital | 3 (1.3) | 3 (1.8) | 0 (0.0) |  |  |
| Bungoma County Referral Hospital | 74 (33.0) | 46 (27.5) | 28 (49.1) |  |  |
| Cheptais Sub-County Hospital | 9 (4.0) | 9 (5.4) | 0 (0.0) |  |  |
| Kabula Health Centre | 14 (6.2) | 14 (8.4) | 0 (0.0) |  |  |
| Kimilili Sub-County Hospital | 48 (21.4) | 40 (24.0) | 8 (14.0) |  |  |
| Naitiri Sub-County Hospital | 6 (2.7) | 6 (3.6) | 0 (0.0) |  |  |
| Sirisia Sub-County Hospital | 2 (0.9) | 2 (1.2) | 0 (0.0) |  |  |
| Webuye County Hospital | 68 (30.4) | 47 (28.1) | 21 (36.8) |  |  |

*Continuous variables are median (interquartile range); categorical variables are n (%). SMD, standardised mean difference between newborns discharged alive and newborns who died; values above 0.10 conventionally indicate imbalance. Percentages use the column denominator. Missing n is the number of mortality-cohort records in which the value was not recorded in the source record. Values not recorded were completed during the independent data audit for maternal age, antenatal visits, multiple pregnancy, hypertensive disorder, anaemia, 5-minute Apgar score and admission conditions, and for antenatal corticosteroids among newborns of 24–34 weeks' gestation; summaries include these completed values. Newborn sex was not recorded in the abstraction instrument*

Condition at admission showed the same gradient. Respiratory distress was documented in 56 of 57 newborns who died (98.2%) against 65 of 167 discharged alive (38.9%), a standardised difference of 1.66. Hypothermia was present in 63.2% against 21.0%, hypoglycaemia in 33.3% against 13.2% and suspected sepsis in 33.3% against 15.0%. Maternal characteristics were comparatively balanced: maternal age, multiple pregnancy, caesarean delivery, hypertensive disorders, maternal infection and anaemia all had standardised differences below 0.21. Overall, 22.8% of newborns were referred in and 73.2% were inborn.

### Patient-level quality-of-care performance

Documented care was high on average and unevenly distributed across domains (Table 2). Among the 268 newborns in the quality cohort, the median overall PLQS was 87.5% (IQR 77.0–100), the median early-care PLQS 93.5% (IQR 83.3–100) and the median continuing-care PLQS 87.5% (IQR 65.3–100). The early-care score was strongly left-skewed with a pronounced ceiling: 85 of 224 newborns in the mortality cohort (37.9%) scored exactly 100% (Fig 2).

**Fig 2.**
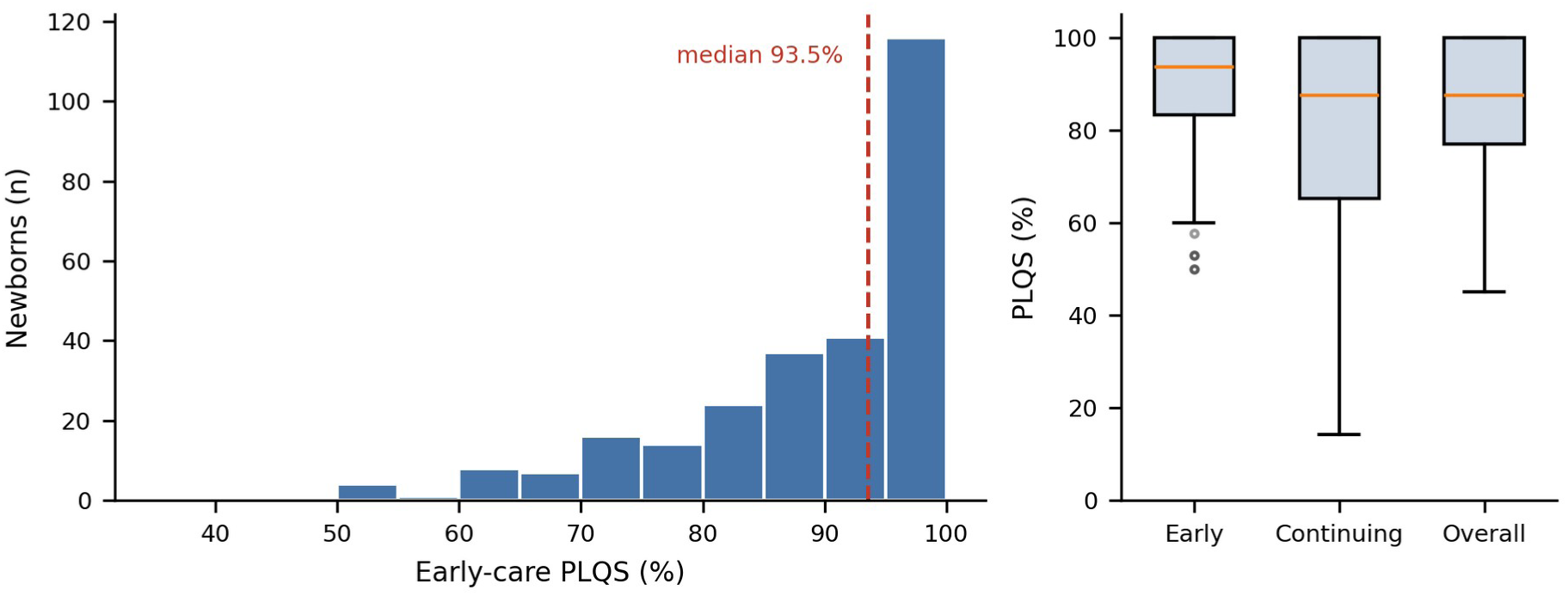
Distribution of the early-care patient-level quality score. Left panel: histogram of the early-care PLQS in the quality-of-care cohort (n=268), with the median marked. Right panel: box plots comparing the early-care, continuing-care and overall PLQS in the same cohort. Boxes show the interquartile range and median; whiskers extend to 1.5 times the interquartile range; points beyond are shown individually

**Table 2.**
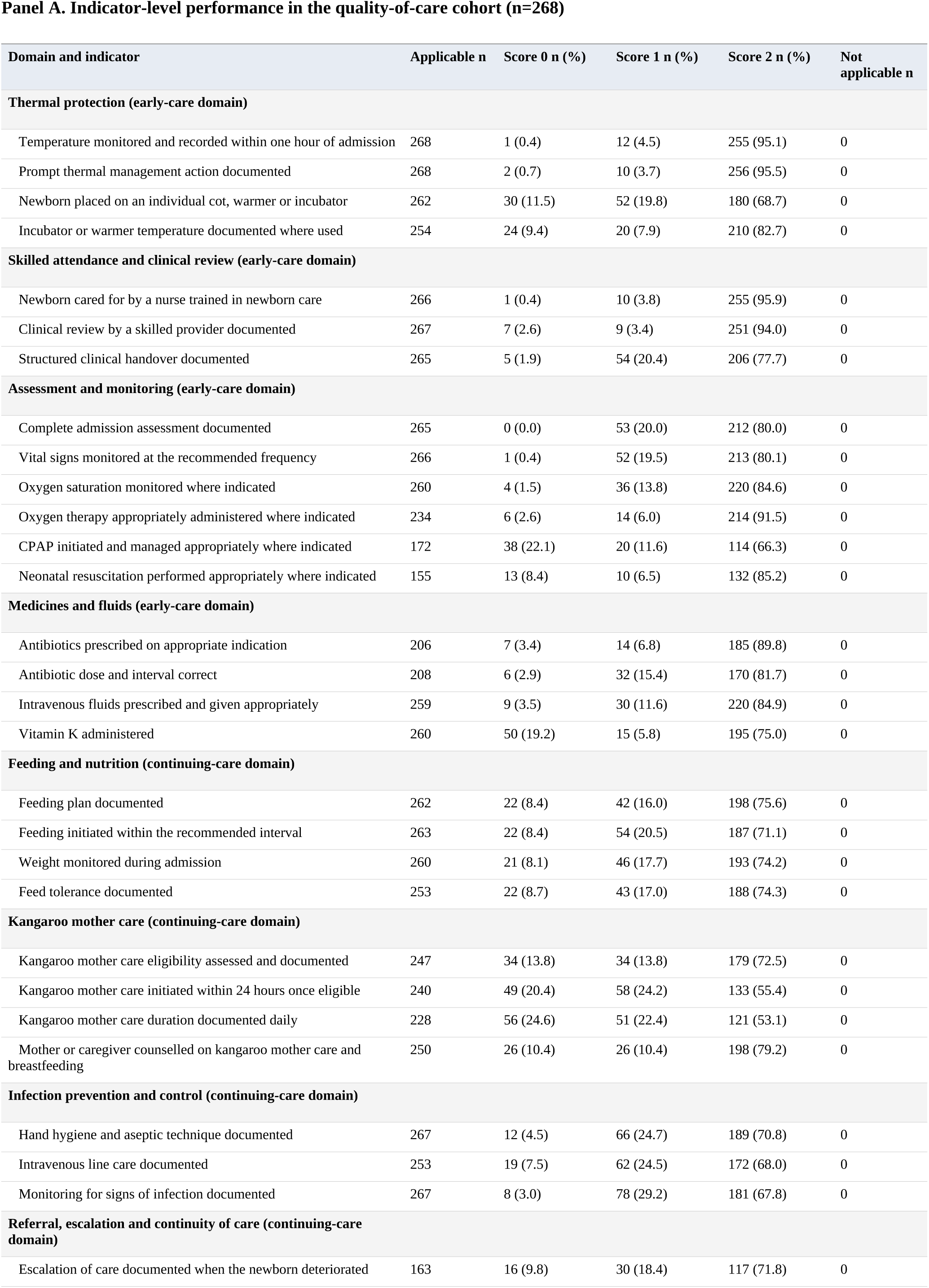
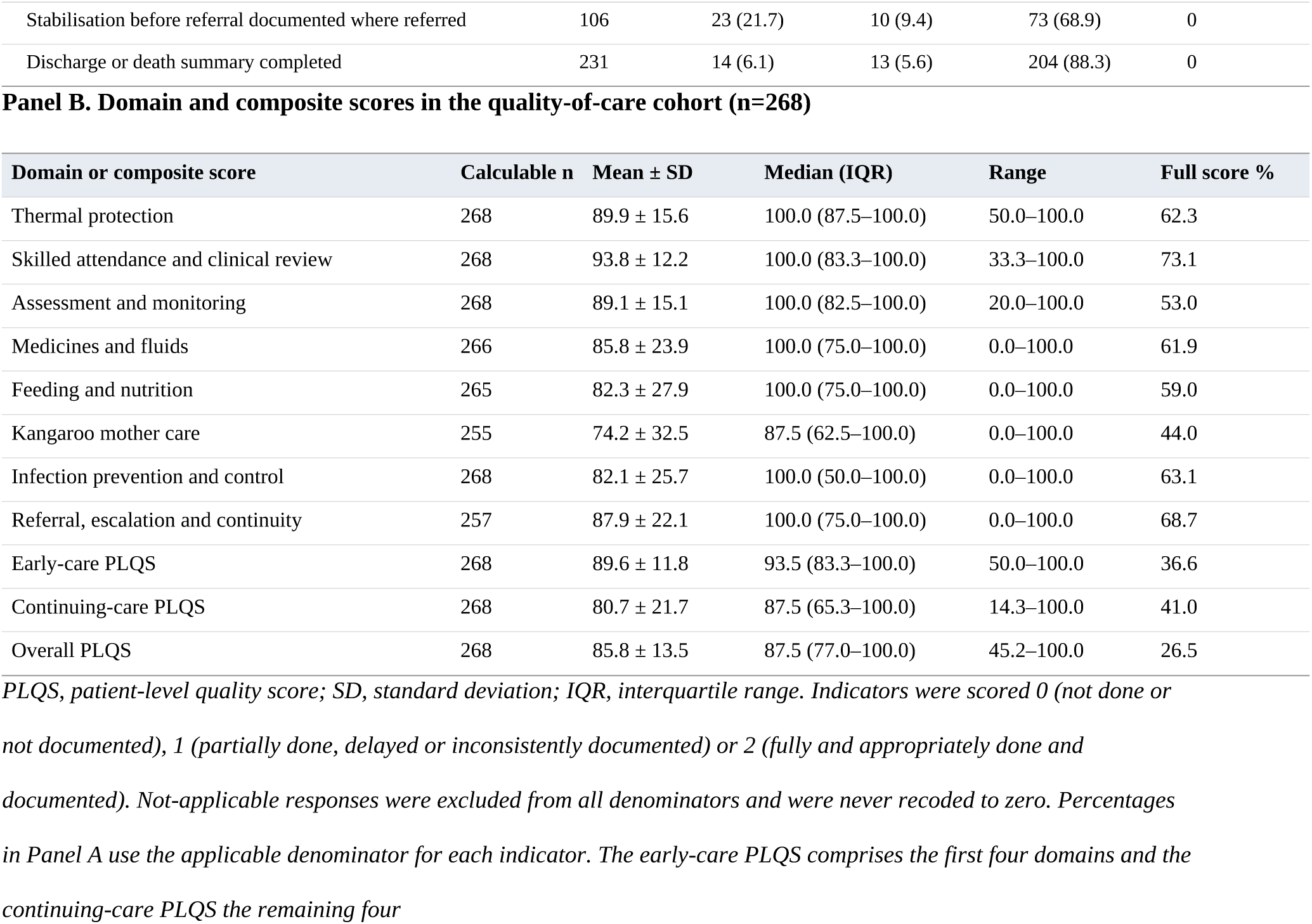
Patient-level quality-of-care performance across PLQS items and domains.

| **Panel A. Indicator-level performance in the quality-of-care cohort (n=268)** |  |  |  |  |  |
|---|---|---|---|---|---|
| **Domain and indicator** | **Applicable n** | **Score 0 n (%)** | **Score 1 n (%)** | **Score 2 n (%)** | **Not applicable n** |
| **Thermal protection (early-care domain)** |  |  |  |  |  |
| Temperature monitored and recorded within one hour of admission | 268 | 1 (0.4) | 12 (4.5) | 255 (95.1) | 0 |
| Prompt thermal management action documented | 268 | 2 (0.7) | 10 (3.7) | 256 (95.5) | 0 |
| Newborn placed on an individual cot, warmer or incubator | 262 | 30 (11.5) | 52 (19.8) | 180 (68.7) | 0 |
| Incubator or warmer temperature documented where used | 254 | 24 (9.4) | 20 (7.9) | 210 (82.7) | 0 |
| **Skilled attendance and clinical review (early-care domain)** |  |  |  |  |  |
| Newborn cared for by a nurse trained in newborn care | 266 | 1 (0.4) | 10 (3.8) | 255 (95.9) | 0 |
| Clinical review by a skilled provider documented | 267 | 7 (2.6) | 9 (3.4) | 251 (94.0) | 0 |
| Structured clinical handover documented | 265 | 5 (1.9) | 54 (20.4) | 206 (77.7) | 0 |
| **Assessment and monitoring (early-care domain)** |  |  |  |  |  |
| Complete admission assessment documented | 265 | 0 (0.0) | 53 (20.0) | 212 (80.0) | 0 |
| Vital signs monitored at the recommended frequency | 266 | 1 (0.4) | 52 (19.5) | 213 (80.1) | 0 |
| Oxygen saturation monitored where indicated | 260 | 4 (1.5) | 36 (13.8) | 220 (84.6) | 0 |
| Oxygen therapy appropriately administered where indicated | 234 | 6 (2.6) | 14 (6.0) | 214 (91.5) | 0 |
| CPAP initiated and managed appropriately where indicated | 172 | 38 (22.1) | 20 (11.6) | 114 (66.3) | 0 |
| Neonatal resuscitation performed appropriately where indicated | 155 | 13 (8.4) | 10 (6.5) | 132 (85.2) | 0 |
| **Medicines and fluids (early-care domain)** |  |  |  |  |  |
| Antibiotics prescribed on appropriate indication | 206 | 7 (3.4) | 14 (6.8) | 185 (89.8) | 0 |
| Antibiotic dose and interval correct | 208 | 6 (2.9) | 32 (15.4) | 170 (81.7) | 0 |
| Intravenous fluids prescribed and given appropriately | 259 | 9 (3.5) | 30 (11.6) | 220 (84.9) | 0 |
| Vitamin K administered | 260 | 50 (19.2) | 15 (5.8) | 195 (75.0) | 0 |
| **Feeding and nutrition (continuing-care domain)** |  |  |  |  |  |
| Feeding plan documented | 262 | 22 (8.4) | 42 (16.0) | 198 (75.6) | 0 |
| Feeding initiated within the recommended interval | 263 | 22 (8.4) | 54 (20.5) | 187 (71.1) | 0 |
| Weight monitored during admission | 260 | 21 (8.1) | 46 (17.7) | 193 (74.2) | 0 |
| Feed tolerance documented | 253 | 22 (8.7) | 43 (17.0) | 188 (74.3) | 0 |
| **Kangaroo mother care (continuing-care domain)** |  |  |  |  |  |
| Kangaroo mother care eligibility assessed and documented | 247 | 34 (13.8) | 34 (13.8) | 179 (72.5) | 0 |
| Kangaroo mother care initiated within 24 hours once eligible | 240 | 49 (20.4) | 58 (24.2) | 133 (55.4) | 0 |
| Kangaroo mother care duration documented daily | 228 | 56 (24.6) | 51 (22.4) | 121 (53.1) | 0 |
| Mother or caregiver counselled on kangaroo mother care and breastfeeding | 250 | 26 (10.4) | 26 (10.4) | 198 (79.2) | 0 |
| **Infection prevention and control (continuing-care domain)** |  |  |  |  |  |
| Hand hygiene and aseptic technique documented | 267 | 12 (4.5) | 66 (24.7) | 189 (70.8) | 0 |
| Intravenous line care documented | 253 | 19 (7.5) | 62 (24.5) | 172 (68.0) | 0 |
| Monitoring for signs of infection documented | 267 | 8 (3.0) | 78 (29.2) | 181 (67.8) | 0 |
| **Referral, escalation and continuity of care (continuing-care domain)** |  |  |  |  |  |
| Escalation of care documented when the newborn deteriorated | 163 | 16 (9.8) | 30 (18.4) | 117 (71.8) | 0 |
| Stabilisation before referral documented where referred | 106 | 23 (21.7) | 10 (9.4) | 73 (68.9) | 0 |
| Discharge or death summary completed | 231 | 14 (6.1) | 13 (5.6) | 204 (88.3) | 0 |
| **Panel B. Domain and composite scores in the quality-of-care cohort (n=268)** |  |  |  |  |  |
| **Domain or composite score** | **Calculable n** | **Mean ± SD** | **Median (IQR)** | **Range** | **Full score %** |
| Thermal protection | 268 | 89.9 ± 15.6 | 100.0 (87.5–100.0) | 50.0–100.0 | 62.3 |
| Skilled attendance and clinical review | 268 | 93.8 ± 12.2 | 100.0 (83.3–100.0) | 33.3–100.0 | 73.1 |
| Assessment and monitoring | 268 | 89.1 ± 15.1 | 100.0 (82.5–100.0) | 20.0–100.0 | 53.0 |
| Medicines and fluids | 266 | 85.8 ± 23.9 | 100.0 (75.0–100.0) | 0.0–100.0 | 61.9 |
| Feeding and nutrition | 265 | 82.3 ± 27.9 | 100.0 (75.0–100.0) | 0.0–100.0 | 59.0 |
| Kangaroo mother care | 255 | 74.2 ± 32.5 | 87.5 (62.5–100.0) | 0.0–100.0 | 44.0 |
| Infection prevention and control | 268 | 82.1 ± 25.7 | 100.0 (50.0–100.0) | 0.0–100.0 | 63.1 |
| Referral, escalation and continuity | 257 | 87.9 ± 22.1 | 100.0 (75.0–100.0) | 0.0–100.0 | 68.7 |
| Early-care PLQS | 268 | 89.6 ± 11.8 | 93.5 (83.3–100.0) | 50.0–100.0 | 36.6 |
| Continuing-care PLQS | 268 | 80.7 ± 21.7 | 87.5 (65.3–100.0) | 14.3–100.0 | 41.0 |
| Overall PLQS | 268 | 85.8 ± 13.5 | 87.5 (77.0–100.0) | 45.2–100.0 | 26.5 |

*PLQS, patient-level quality score; SD, standard deviation; IQR, interquartile range. Indicators were scored 0 (not done or not documented), 1 (partially done, delayed or inconsistently documented) or 2 (fully and appropriately done and documented). Not-applicable responses were excluded from all denominators and were never recoded to zero. Percentages in Panel A use the applicable denominator for each indicator. The early-care PLQS comprises the first four domains and the continuing-care PLQS the remaining four*

At domain level, skilled attendance and clinical review scored highest (median 100%, mean 93.8%, 73.1% of newborns achieving a full score) and kangaroo mother care lowest (median 87.5%, mean 74.2%, 44.0% full score) (Fig 3). Between these, thermal protection (mean 89.9%), assessment and monitoring (mean 89.1%), referral and continuity (mean 87.9%), medicines and fluids (mean 85.8%), feeding and nutrition (mean 82.3%) and infection prevention and control (mean 82.1%) formed a broadly similar band, although the last of these had the widest lower interquartile bound of any domain at 50%.

**Fig 3.**
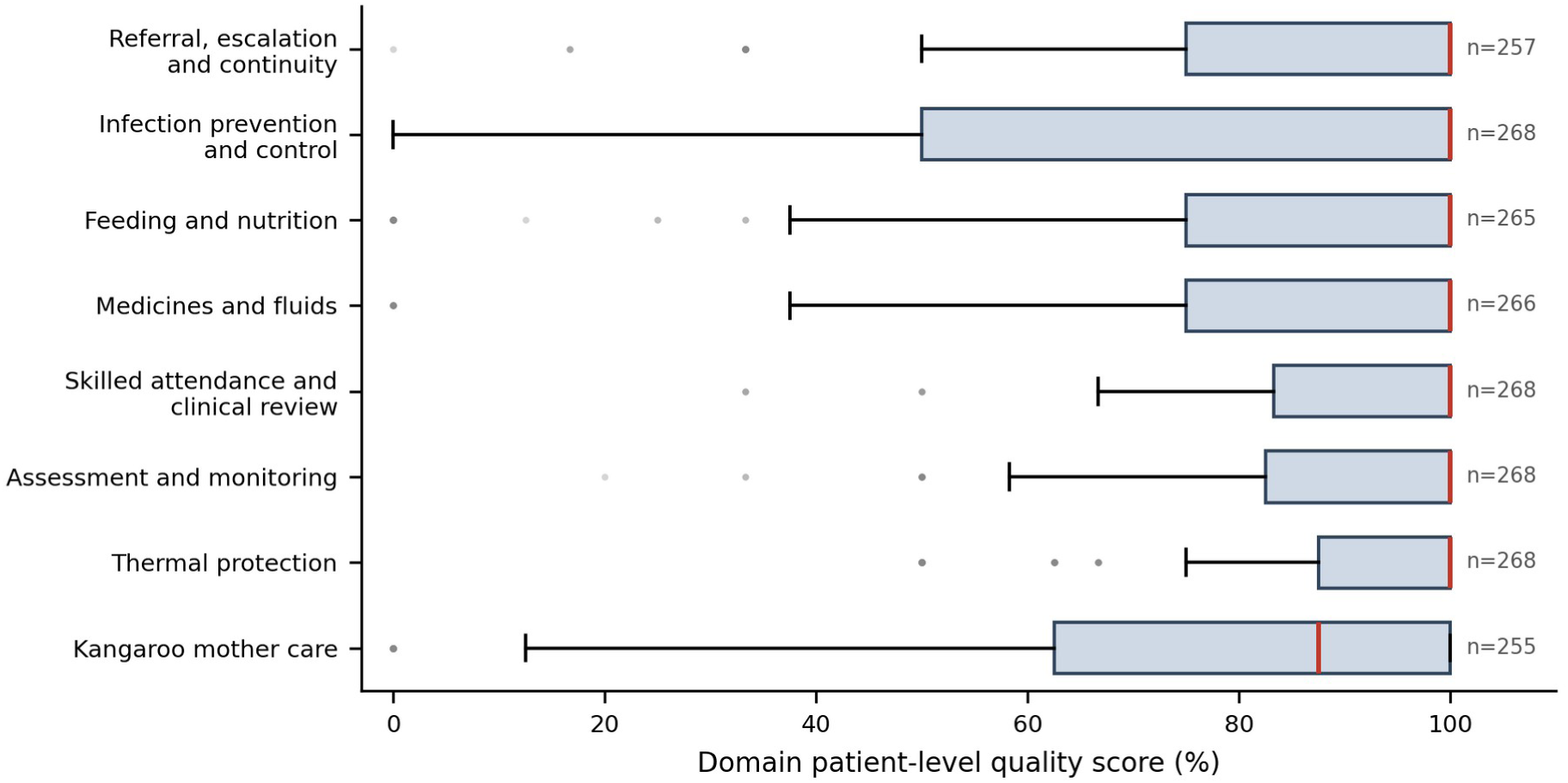
Domain-level patient-level quality-of-care profile. Box plots of the eight PLQS domain scores in the quality-of-care cohort, ordered by median. The number of newborns for whom each domain was calculable is shown to the right of each box. Boxes show the interquartile range, the red line the median, whiskers 1.5 times the interquartile range, and points beyond are shown individually

Item-level performance identified specific deficits that domain averages concealed. Within the early-care domains, temperature monitoring within one hour of admission was fully documented in 255 of 268 applicable records (95.1%) and care by a nurse trained in newborn care in 255 of 266 (95.9%), but placing the newborn on an individual cot, warmer or incubator was fully documented in only 180 of 262 (68.7%), with 30 records (11.5%) scoring zero. CPAP was initiated and managed appropriately in 114 of 172 applicable records (66.3%), the lowest early-care item, with 38 (22.1%) scoring zero. Vitamin K administration was fully documented in 195 of 260 (75.0%) with 50 (19.2%) scoring zero. Within the continuing-care domains, kangaroo mother care initiation within 24 hours of eligibility was fully documented in 133 of 240 applicable records (55.4%) and daily duration in 121 of 228 (53.1%), the two weakest indicators in the instrument. Stabilisation before referral was fully documented in 73 of 106 applicable records (68.9%), with 23 (21.7%) scoring zero.

### In-hospital outcomes

In-hospital mortality in the mortality cohort was 25.4% (57 of 224). Complications were documented during admission in 142 of 265 newborns in the quality cohort (53.6%) and sepsis in 67 (25.3%). Among the 167 discharged survivors, median length of stay was 8.2 days (IQR 4.1–16.9) and human-milk feeding at discharge was recorded for 149 of 163 with a documented feeding mode (91.4%).

Outcomes across early-care PLQS categories showed no monotonic gradient (Table 3). Mortality was 25.0% in the lowest category (≤85.3%), 36.5% in the second (85.4–94.1%), 55.6% in the third (94.2–99.9%) and 9.4% among newborns scoring exactly 100%. Median gestational age was 34 weeks in the lowest category and 32 weeks in the second, indicating that the categories differ substantially in baseline risk and cannot be compared without adjustment.

**Table 3.**
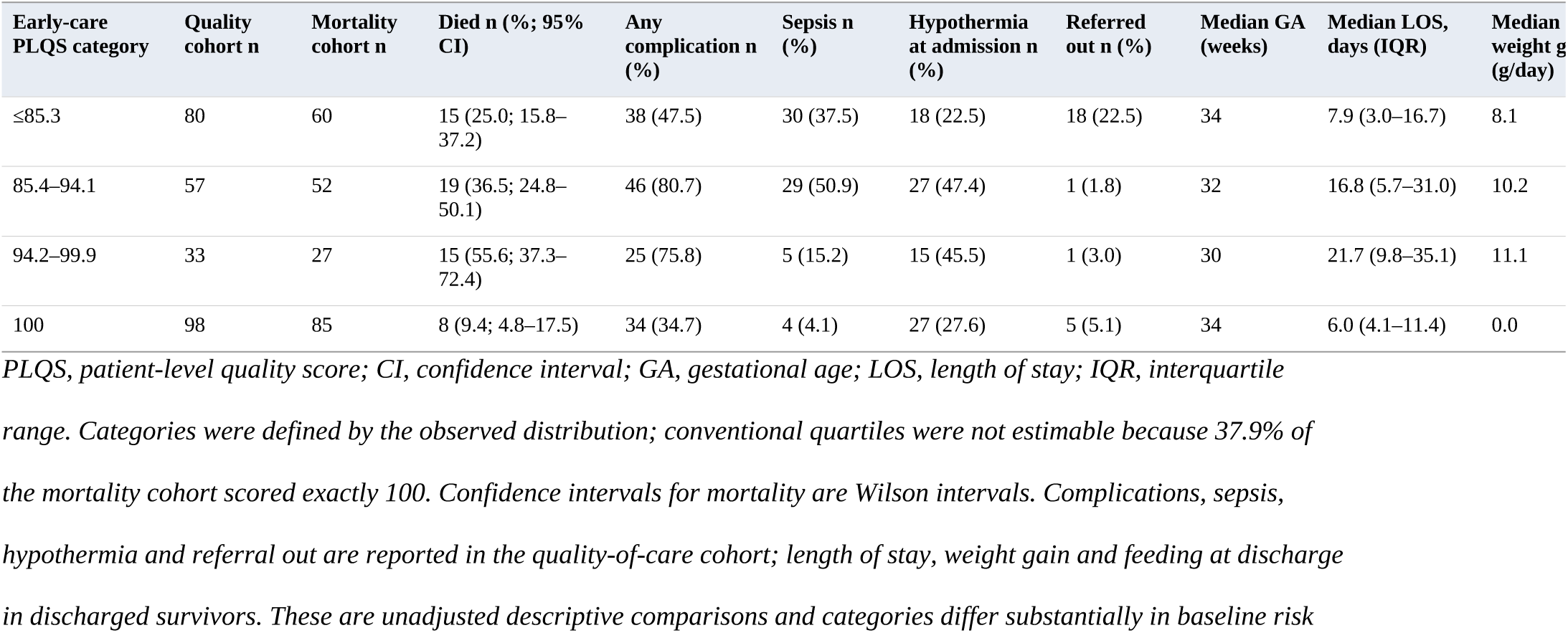
Clinical outcomes according to early-care PLQS category.

| Early-care PLQS category | Quality cohort n | Mortality cohort n | Died n (%; 95% CI) | Any complication n (%) | Sepsis n (%) | Hypothermia at admission n (%) | Referred out n (%) | Median GA (weeks) | Median LOS, days (IQR) | Median weight g[illegible] (g/day) |
|---|---|---|---|---|---|---|---|---|---|---|
| ≤85.3 | 80 | 60 | 15 (25.0; 15.8–37.2) | 38 (47.5) | 30 (37.5) | 18 (22.5) | 18 (22.5) | 34 | 7.9 (3.0–16.7) | 8.1 |
| 85.4–94.1 | 57 | 52 | 19 (36.5; 24.8–50.1) | 46 (80.7) | 29 (50.9) | 27 (47.4) | 1 (1.8) | 32 | 16.8 (5.7–31.0) | 10.2 |
| 94.2–99.9 | 33 | 27 | 15 (55.6; 37.3–72.4) | 25 (75.8) | 5 (15.2) | 15 (45.5) | 1 (3.0) | 30 | 21.7 (9.8–35.1) | 11.1 |
| 100 | 98 | 85 | 8 (9.4; 4.8–17.5) | 34 (34.7) | 4 (4.1) | 27 (27.6) | 5 (5.1) | 34 | 6.0 (4.1–11.4) | 0.0 |

*PLQS, patient-level quality score; CI, confidence interval; GA, gestational age; LOS, length of stay; IQR, interquartile range. Categories were defined by the observed distribution; conventional quartiles were not estimable because 37.9% of the mortality cohort scored exactly 100. Confidence intervals for mortality are Wilson intervals. Complications, sepsis, hypothermia and referral out are reported in the quality-of-care cohort; length of stay, weight gain and feeding at discharge in discharged survivors. These are unadjusted descriptive comparisons and categories differ substantially in baseline risk*

### Early-care quality of care and in-hospital mortality

There was no evidence of an association between documented early-care quality and in-hospital mortality in any model (Table 4). Unadjusted, the risk ratio per 10-percentage-point increase in early-care PLQS was 0.98 (95% CI 0.84–1.15, p=0.821). After adjustment for gestational age, multiple pregnancy, birth location and facility it was 1.06 (95% CI 0.85– 1.31, p=0.627), and after additional adjustment for 5-minute Apgar score, hypothermia at admission and suspected sepsis at admission it was 1.03 (95% CI 0.82–1.29, p=0.797). The confidence intervals exclude effects larger than about a 20% reduction or a 30% increase in risk per 10-point increment.

**Table 4.** Association between early-care patient-level quality of care and in-hospital mortality.

| Exposure or covariate | n | Deaths | Model 1 RR (95% CI) | p | Model 2 aRR (95% CI) | p | Model 3 aRR (95% CI) | p |
|---|---|---|---|---|---|---|---|---|
| Early-care PLQS, per 10-percentage-point increase | 224 | 57 | 0.98 (0.84–1.15) | 0.821 | 1.05 (0.85–1.31) | 0.627 | 1.03 (0.82–1.29) | 0.797 |
| **Early-care PLQS category** |  |  |  |  |  |  |  |  |
| ≤85.3% | 60 | 15 |  |  |  |  | 1.00 (reference) |  |
| 85.4–94.1% | 52 | 19 |  |  |  |  | 0.97 (0.62–1.49) | 0.878 |
| 94.2–99.9% | 27 | 15 |  |  |  |  | 1.90 (1.08–3.37) | 0.027 |
| 100% | 85 | 8 |  |  |  |  | 0.64 (0.32–1.30) | 0.222 |
| **Other covariates in Model 3** |  |  |  |  |  |  |  |  |
| Gestational age, per completed week |  |  |  |  |  |  | 0.83 (0.77–0.89) | <0.001 |
| Multiple pregnancy |  |  |  |  |  |  | 1.02 (0.72–1.44) | 0.921 |
| Outborn |  |  |  |  |  |  | 0.89 (0.54–1.46) | 0.636 |
| Referral hospital with newborn unit |  |  |  |  |  |  | 2.00 (1.00–3.99) | 0.051 |
| 5-minute Apgar, per point |  |  |  |  |  |  | 0.92 (0.81–1.04) | 0.171 |
| Hypothermia at admission |  |  |  |  |  |  | 1.80 (1.09–2.97) | 0.022 |
| Suspected sepsis at admission |  |  |  |  |  |  | 1.67 (0.97–2.89) | 0.065 |

*RR, risk ratio; aRR, adjusted risk ratio; CI, confidence interval; PLQS, patient-level quality score. All estimates are from modified Poisson regression with robust (sandwich) variance in the mortality cohort (n=224, 57 deaths). Model 1 is unadjusted. Model 2 is adjusted for gestational age, multiple pregnancy, birth location (inborn or outborn) and facility. Model 3 is additionally adjusted for 5-minute Apgar score, hypothermia at admission and suspected sepsis at admission. Facility was modelled as a binary term (the two referral hospitals with operational newborn units against all other facilities) because individual facility fixed effects produced complete separation in the five facilities recording no deaths. Respiratory distress at admission was excluded from Model 3 because of near-complete separation; its crude risk ratio was 47.7 (95% CI 6.7–338.4) and its adjusted risk ratio when forced into Model 3 was 23.5 (95% CI 3.3–166.5). Covariates not recorded in the source records were completed during the independent data audit (S1 Table). Gestational age was missing in 3 of 224 records (1.3%) and was handled by 20 imputations by chained equations with Rubin's rules pooling; the complete-case estimate for the primary exposure in Model 3 was 1.02 (95% CI 0.81–1.27). Category estimates are from Model 3 with the lowest category as reference*

Baseline maturity and admission condition dominated the models. Each additional completed week of gestation was associated with a 17% lower risk of death (adjusted risk ratio 0.83, 95% CI 0.77–0.89, p<0.001), hypothermia at admission with a 1.80-fold higher risk (95% CI 1.09–2.97, p=0.022) and suspected sepsis at admission with a 1.67-fold higher risk (95% CI 0.97–2.89, p=0.065). Respiratory distress at admission, excluded from the primary model because of near-complete separation, had a crude risk ratio of 47.7 (95% CI 6.7–338.4) and an adjusted risk ratio of 23.5 (95% CI 3.3–166.5) when forced into Model 3; the extreme width of these intervals reflects that only 1 of 103 newborns without documented respiratory distress died, and these estimates should be read as indicating a strong but imprecisely quantified association.

Modelled in categories, the adjusted risk ratio relative to the lowest category was 0.97 (95% CI 0.63–1.50) for 85.4–94.1%, 1.91 (95% CI 1.08–3.37) for 94.2–99.9% and 0.64 (95% CI 0.32–1.31) for a score of exactly 100% (Fig 4). This non-monotonic pattern, in which the second-highest category carried the highest adjusted risk, is not consistent with a dose– response relationship. Neither a quadratic term (risk ratio 0.91, 95% CI 0.66–1.25, p=0.555) nor a restricted cubic spline term (p=0.301) provided evidence of nonlinearity, so a spline figure was not fitted.

**Fig 4.**
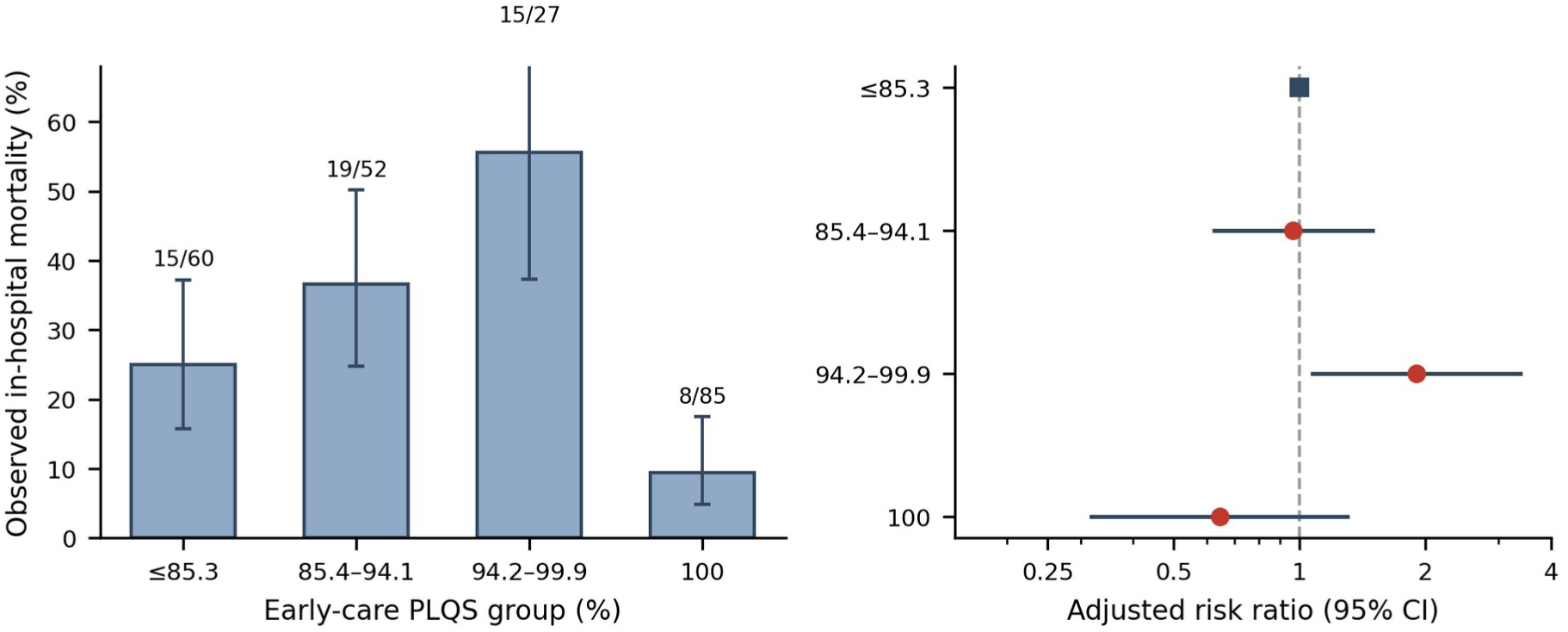
Early-care patient-level quality score and in-hospital mortality. Left panel: observed in-hospital mortality by early-care PLQS category in the mortality cohort (n=224), with Wilson 95% confidence intervals and the number of deaths over the number of newborns shown above each bar. Right panel: adjusted risk ratios with 95% confidence intervals from a modified Poisson model adjusted for gestational age, multiple pregnancy, birth location, facility, 5-minute Apgar score, hypothermia at admission and suspected sepsis at admission, with the lowest category as reference. The horizontal axis is on a logarithmic scale

### Secondary clinical outcomes

Higher early-care PLQS was not associated with any complication during admission (adjusted risk ratio 1.01 per 10 points, 95% CI 0.91–1.11), with hypothermia during admission (0.95, 95% CI 0.69–1.32), with discharge alive (1.01, 95% CI 0.97–1.06), with length of stay (adjusted ratio of means 1.07, 95% CI 0.86–1.31), with weight change (adjusted mean difference −17.5 g, 95% CI −40.8 to 5.8) or with average daily weight gain (−1.6 g/day, 95% CI −4.8 to 1.5) (Table 5; full covariate output in S5 Table; model diagnostics in S6 Table).

**Table 5.**
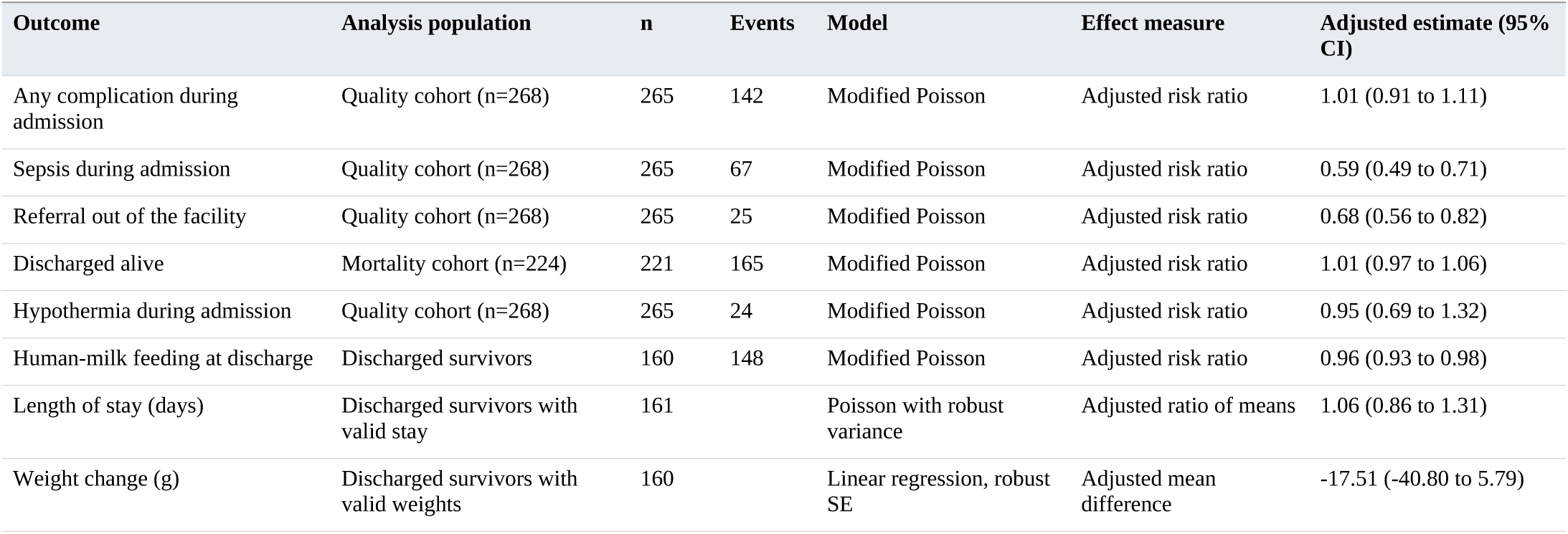
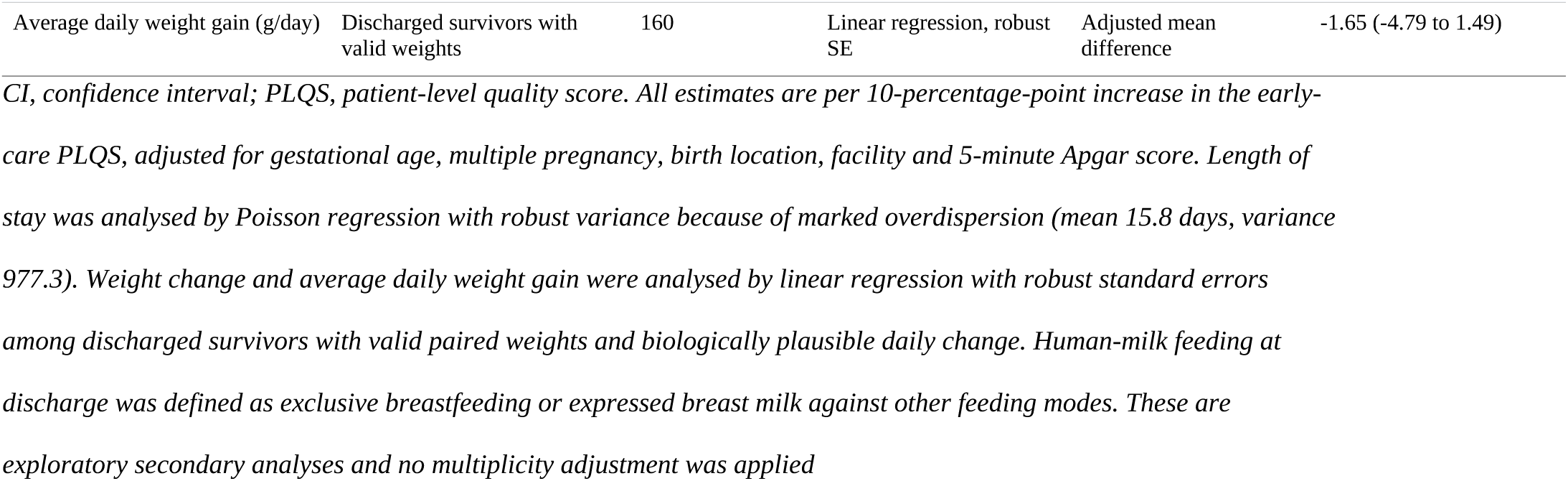
Associations between early-care quality of care and secondary newborn outcomes.

Three secondary associations were inverse and had confidence intervals excluding the null. Higher early-care PLQS was associated with lower documented sepsis during admission (adjusted risk ratio 0.59 per 10 points, 95% CI 0.49–0.72, p<0.001), with lower referral out of the facility (0.68, 95% CI 0.56–0.82, p<0.001) and, unexpectedly, with marginally lower human-milk feeding at discharge (0.96, 95% CI 0.93–0.99, p=0.003). Given that these are exploratory analyses among ten secondary outcomes, and that documented sepsis and documented care are both products of the same clinical record, these findings are interpreted cautiously below.

### Sensitivity and bias-control analyses

The null primary finding was robust (Table 6; S3 and S4 Tables). Complete-case analysis for gestational age with the Model 3 covariate set gave an adjusted risk ratio of 1.02 (95% CI 0.81–1.27) against 1.03 in the pooled analysis. Substituting birth-weight category for gestational age gave 0.94 (95% CI 0.75–1.17). Restriction to the 24-hour survivor cohort gave 1.03 (95% CI 0.80–1.32), and restriction to newborns with at least three of four early-care domains valid gave 1.02 (95% CI 0.81–1.27). Stratified analyses gave 1.09 (95% CI 0.87–1.37) among 163 inborn newborns and 0.65 (95% CI 0.38–1.12) among 58 outborn newborns; the confidence interval in the smaller outborn stratum is wide and includes the null. Restricting the analysis to the 181 newborns with every Model 3 covariate recorded in the source records gave 1.05 (95% CI 0.80–1.36), and pooling across the 20 completed covariate datasets gave 0.99 (95% CI 0.79–1.25).

**Table 6.** Sensitivity and bias-control analyses: early-care PLQS per 10-percentage-point increase and in-.

| **Analysis** | **n** | **Deaths** | **Adjusted risk ratio (95% CI)** | **p** |
|---|---|---|---|---|
| Complete case for gestational age, Model 2 covariates | 221 | 56 | 1.05 (0.84–1.30) | 0.688 |
| Complete case for gestational age, Model 3 covariates | 221 | 56 | 1.02 (0.81–1.27) | 0.884 |
| Complete case on source-record covariates (Model 3) | 181 | 48 | 1.05 (0.80–1.36) | 0.742 |
| Pooled across 20 completed covariate datasets (Model 3) | 221 | 56 | 0.99 (0.79–1.25) | 0.953 |
| Birth-weight category replacing gestational age (Model 3) | 223 | 57 | 0.94 (0.75–1.17) | 0.564 |
| 24-hour survivor cohort (Model 3) | 215 | 50 | 1.03 (0.80–1.32) | 0.824 |
| Restricted to ≥3 of 4 early-care domains valid (Model 3) | 221 | 56 | 1.02 (0.81–1.27) | 0.884 |
| Inborn newborns only (Model 3 without inborn term) | 163 | 37 | 1.09 (0.87–1.37) | 0.448 |
| Outborn newborns only (Model 3 without inborn term) | 58 | 19 | 0.65 (0.38–1.12) | 0.119 |
| Overall PLQS per 10 points (Model 3) | 221 | 56 | 0.77 (0.66–0.90) | 0.001 |
| Continuing-care PLQS per 10 points (Model 3) | 221 | 56 | 0.81 (0.74–0.89) | <0.001 |

*CI, confidence interval; PLQS, patient-level quality score. Unless otherwise stated, models are adjusted as for Model 3. The final two rows substitute alternative exposure definitions into the identical Model 3 specification in the same population and are the analyses referred to in the text as demonstrating opportunity-to-receive-care bias. Source-record complete case excludes newborns with any Model 3 covariate not recorded in the source records; pooled estimates combine the 20 completed covariate datasets by Rubin's rules, with gestational age analysed as complete case*

Neither interaction was supported. The interaction between early-care PLQS and early preterm status below 32 weeks gave a ratio of risk ratios of 1.09 (95% CI 0.66–1.80, p=0.735), with stratum-specific estimates of 1.01 among newborns of 32 weeks or more and 1.05 among those below 32 weeks. The interaction with referred-in status gave 1.00 (95% CI 0.56–1.79, p=0.998). Both interaction analyses are underpowered and are reported as inconclusive.

The most informative sensitivity analysis concerned the exposure definition itself. Replacing the early-care PLQS with the overall PLQS in the identical Model 3 specification produced an apparently strong inverse association (adjusted risk ratio 0.77 per 10 points, 95% CI 0.66– 0.90, p=0.001), and replacing it with the continuing-care PLQS produced a stronger one still (0.81, 95% CI 0.74–0.89, p<0.001). The exposure definition therefore determined the direction and strength of the finding while the outcome, covariates, model and population were held constant.

### Facility variation

Median early-care PLQS varied across the facilities contributing at least five newborns, and case mix varied with it (Fig 5; S2 Table). Facility variation is presented descriptively only: the two referral hospitals with newborn units contributed 142 of 224 newborns in the mortality cohort and 49 of 57 deaths, and receive the smallest and sickest newborns in the network, so facility differences in documented quality cannot be separated from differences in case mix in these data. In the adjusted mortality models, care at a referral hospital with a newborn unit was associated with a higher risk of death (adjusted risk ratio 2.00, 95% CI 1.00–3.99 in Model 3), which is consistent with referral of higher-risk newborns rather than with any inference about facility performance.

**Fig 5.**
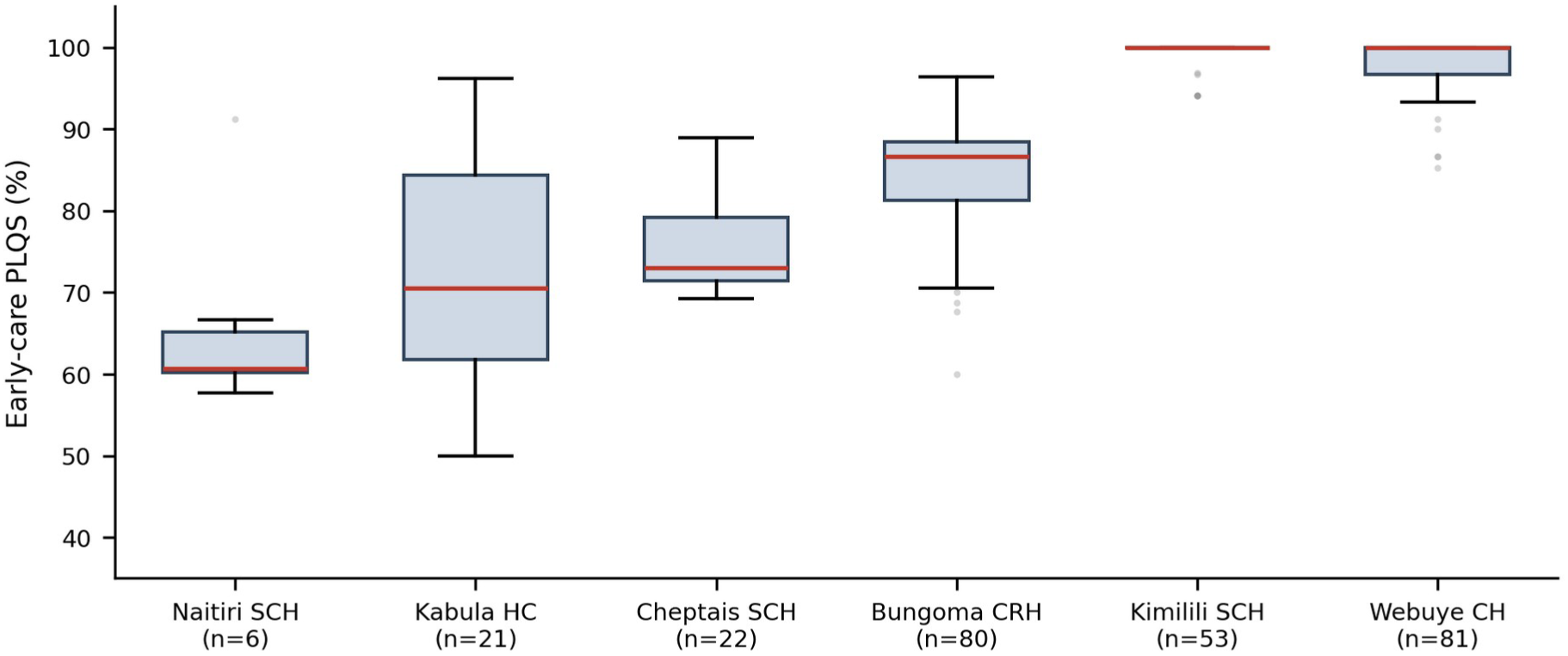
Facility variation in the early-care patient-level quality score. Box plots of the early-care PLQS by facility among facilities contributing at least five newborns to the quality-of-care cohort, ordered by median, with the number of newborns per facility shown on the axis. Facility case mix differs substantially and these distributions should not be read as a comparison of facility performance. CRH, county referral hospital; CH, county hospital; SCH, sub-county hospital; HC, health centre

## Discussion

### Principal findings

Among 268 preterm and low-birth-weight newborns admitted to eight facilities in Western Kenya, documented early patient-level quality of care was high (median early-care PLQS 93.5%) but was not associated with in-hospital mortality. The adjusted risk ratio was 1.03 per 10-percentage-point increase (95% CI 0.82–1.29), and this null result persisted across complete-case analysis, birth-weight substitution, restriction to 24-hour survivors, restriction to records with at least three valid early-care domains, and stratification by birth location. Mortality was instead dominated by maturity and condition at admission: each additional week of gestation was associated with a 17% lower risk of death, hypothermia at admission with a 1.80-fold higher risk, and respiratory distress at admission with an association so strong that it could not be stably estimated.

The single most consequential finding is methodological. Holding outcome, covariates, model and population constant and changing only the exposure definition reversed the result: the continuing-care score appeared strongly protective (adjusted risk ratio 0.81 per 10 points, 95% CI 0.74–0.89) where the early-care score showed nothing. This is a direct empirical demonstration, within one dataset, of the magnitude of opportunity-to-receive-care bias in composite quality scores.

### Interpreting the null association between early care and mortality

A null finding here should not be read as evidence that quality of care does not matter for preterm survival. Three explanations are more plausible and are not mutually exclusive.

First, the exposure had little room to vary. The early-care score was strongly left-skewed, with 37.9% of newborns scoring exactly 100% and an interquartile range of 83.3–100. Where a process measure approaches its ceiling in most newborns, the contrast available to detect an association is small and largely confined to the tail of the distribution. Documented adherence to the early-care processes captured here was, in these facilities, close to complete for most newborns; the variation that remained may reflect documentation practice as much as care delivery.

Second, the score measures whether prespecified processes were documented, not whether they were effective in the individual newborn; the general limitations of routine facility data for newborn indicators are well established [14,31]. An indicator scored 2 for appropriate CPAP management records that CPAP was initiated and managed according to protocol, not that the newborn’s respiratory failure was reversed. The interventions most likely to alter survival for a newborn of 27 weeks — surfactant and mechanical ventilation — were not available in these facilities and therefore could not be scored at all. A process score constructed from the care that is available cannot capture the effect of the care that is absent.

Third, confounding by indication runs in the opposite direction to the hypothesis. Sicker newborns attract more intensive documented care: they are more likely to receive CPAP, oxygen, antibiotics and frequent monitoring, and each of these generates a scoreable item. Newborns with the highest documented early care therefore include a disproportionate number of the most compromised. The non-monotonic category pattern we observed, in which adjusted risk was highest in the second-highest score category, is consistent with this. Adjustment for gestational age, Apgar score, hypothermia and suspected sepsis reduces but almost certainly does not eliminate this confounding, because the intensity of illness within a given gestational-age and admission-condition stratum is not fully captured by these variables.

### Opportunity-to-receive-care bias and the choice of exposure

The contrast between the null early-care result and the apparently strong continuing-care result deserves emphasis because it has direct implications for how newborn quality-of-care research is conducted. Sustained kangaroo mother care, daily weight monitoring, feed tolerance documentation and a completed discharge summary can only accrue to a newborn who survives long enough. Where such items enter a composite exposure, the score partly measures survival time, and its association with mortality is partly tautological. Our data quantify this: an exposure that included these items produced an adjusted risk ratio of 0.81 per 10 points, while an exposure restricted to stabilisation-phase processes produced 1.03 in the same 224 newborns with identical adjustment.

This is structurally the same problem as immortal time bias, in which exposure classification depends on surviving a period during which the outcome could occur, and which has been shown to generate substantial spurious protective associations in observational pharmacoepidemiology [15,16]. To our knowledge it has been less often addressed in facility-based newborn quality research, where composite adherence scores spanning the whole admission are common. Restricting the exposure to early-care processes, as we prespecified, is a simple and transparent partial remedy; excluding deaths in the first 24 hours, which we also did, addresses a related but distinct component. Studies reporting protective associations between whole-admission quality scores and neonatal survival should state explicitly which items could only be delivered to survivors.

### Domain-specific quality gaps

Although composite early care was high, specific indicators were not, and these are the actionable findings. Kangaroo mother care was the weakest domain (mean 74.2%), with initiation within 24 hours of eligibility fully documented in 55.4% of applicable records and daily duration in 53.1%. Given that immediate kangaroo mother care reduces mortality by around a quarter among newborns weighing 1,000–1,799 g [20] and that WHO now recommends initiation as soon as possible after birth [21], this is the domain with the clearest evidence-to-practice gap in our data. Barriers identified across sub-Saharan Africa — space, beds, staffing and caregiver availability — are well described [22,23], and multi-country bottleneck analysis identified the same constraints for basic newborn care and resuscitation [28], and our finding that documentation was weakest for sustained rather than initial kangaroo mother care is consistent with capacity rather than intent being the constraint.

Two early-care indicators also stood out. Placing the newborn on an individual cot, warmer or incubator was fully documented in 68.7% of records, with 11.5% scoring zero, indicating that equipment sharing remains routine; this matters because hypothermia at admission was present in 31.7% of our cohort and was independently associated with an 80% higher risk of death, consistent with the association between admission hypothermia and mortality reported across the Kenyan Clinical Information Network [24]. Appropriate CPAP initiation and management was fully documented in 66.3% of applicable records, the weakest early-care indicator, in a setting where regional assessments have found CPAP readiness at around 46% of the achievable maximum in Kenyan neonatal units [8]; trial evidence indicates that investment in oxygen systems rather than devices alone improves outcomes for children and neonates in comparable settings [29]. Vitamin K administration was not documented at all in 19.2% of records, a low-cost omission with a straightforward remedy.

### Secondary outcomes and their interpretation

Three inverse secondary associations require careful reading rather than acceptance. The association between higher early-care PLQS and lower documented sepsis (adjusted risk ratio 0.59 per 10 points) is not readily interpretable as a benefit of care, because both the exposure and the outcome derive from the same clinical record: a newborn with sparse documentation will tend to have both a lower score and fewer documented complications. The association with lower referral out of the facility (0.68) is more plausibly explained by capability — facilities documenting more complete early care are those able to retain and manage newborns rather than transfer them. The small inverse association with human-milk feeding at discharge (0.96) is of uncertain significance among ten exploratory outcomes and, with 91.4% of survivors receiving human milk, has little room to be clinically meaningful. None of these should be presented as evidence that better care produces worse feeding or that care prevents sepsis.

### Comparison with existing evidence

In-hospital mortality of 25.4% among preterm and low-birth-weight newborns in our cohort is high but within the range reported for Kenyan newborn units caring for comparable populations. A multisite retrospective cohort across the Kenyan Clinical Information Network reported substantial between-hospital variation in neonatal inpatient mortality after case-mix adjustment [11], and earlier assessments of neonatal care in Kenyan clinical training facilities documented widespread deficits in essential newborn processes [12]. Our cohort was more severely selected than most: 32.1% were born before 32 weeks and 26.8% weighed under 1,500 g. Kenya’s national progress in reducing under-five mortality has been driven substantially by factors outside inpatient newborn care [36], and facility-based perinatal death review remains inconsistently translated into change across the region [37].

The direction of our principal finding differs from analyses of survey-based newborn care indicators. An analysis of the 2022 Kenya Demographic and Health Survey found that only 32.7% of mothers reported their newborn receiving all components of quality newborn care, and interpreted low coverage as a driver of neonatal mortality [25]. That analysis concerns coverage of basic postnatal processes across the whole birth cohort as reported by mothers; ours concerns documented adherence to inpatient processes among a severely preterm hospitalised population where coverage was already high. The two are not in conflict: where coverage is low, increasing it plausibly reduces mortality, whereas where documented adherence is already near ceiling among the available interventions, further gains depend on capabilities the facilities do not yet have.

Our finding that facility-level factors and case mix dominate is consistent with regional service-readiness evidence. Assessments of 64 neonatal units across Kenya, Malawi, Nigeria and Tanzania found median service readiness for WHO level-2+ care around half of the achievable maximum, with respiratory support the principal gap [8,9], and nurse-to-baby ratios in the same network of 1:6 by day and 1:10 at night [26]. Where surfactant and mechanical ventilation are unavailable and staffing is several times above recommended workload [27], process adherence within the available repertoire may be a weak lever on survival for the smallest newborns.

### Health-system and clinical implications

Three implications follow. First, quality-improvement effort in these facilities should target the indicators that were genuinely deficient — sustained kangaroo mother care, individual thermal support, CPAP management and vitamin K — rather than the composite score, which is already high and would move little. Second, admission hypothermia deserves priority as a modifiable admission-state exposure independently associated with mortality in our data and in larger Kenyan series [24]; it is addressable through thermal capability at the point of admission and during transfer. Third, for the most immature newborns, the constraint appears to lie in the repertoire of available care rather than in adherence to it, which points towards capability investment — respiratory support and the staffing to supervise it — rather than towards adherence campaigns. Newborns transferred between facilities are a particular concern, since neonatal transport across sub-Saharan Africa is poorly resourced and newborns frequently arrive compromised [35].

For research, the implication is that composite quality exposures should be constructed with explicit attention to when each process can occur relative to the outcome. Reporting both an early-care and a whole-admission version, as we have done, makes the magnitude of the bias visible to readers rather than leaving it embedded in the estimate. Structured newborn record platforms of the kind operating in Kenya make such analyses feasible at scale [30,32,33], and standardised measurement definitions for newborn indicators would make them comparable across settings [34].

### Strengths

The study has several strengths. It used an independently audited multicentre newborn dataset in which every raw worksheet was reconciled cell-by-cell against the source export before analysis. It measured quality at the level of the individual newborn across eight care domains and 31 indicators rather than relying on facility structural measures. The primary exposure was prespecified to exclude survival-dependent items, and this decision was tested rather than assumed, by fitting the alternative exposures and reporting the contrast. Relative risks were estimated directly by modified Poisson regression rather than approximated by odds ratios. Nine sensitivity and bias-control analyses, including complete-case, 24-hour survivor and stratified analyses, were prespecified and all are reported. Newborns whose final outcome could not be verified were excluded from the mortality analysis rather than being counted as survivors.

## Limitations

Thirteen limitations qualify these findings. First, the design is a retrospective record review; the exposure and covariates were recorded for clinical rather than research purposes. Second, the PLQS measures documented care, so care delivered but not recorded is scored as not delivered; documentation quality and care quality cannot be separated in these data. Third, and conversely, missing documentation for some indicators may be misclassified as incomplete care. Fourth, residual confounding by disease severity is likely: gestational age, Apgar score, hypothermia and suspected sepsis do not capture the full spectrum of illness intensity within strata. Fifth, confounding by indication operates specifically against the hypothesis, since sicker newborns attract more documented intervention. Sixth, the sample of 268 newborns with 57 deaths limits precision, and the confidence interval around the primary estimate remains compatible with a modest association in either direction. Seventh, newborn sex was not recorded in the abstraction instrument and could not be adjusted for. Eighth, newborns were clustered within eight facilities, and sparse strata required facility to be modelled as a binary term rather than as individual fixed effects. Ninth, gestational age was derived predominantly from last menstrual period rather than early ultrasound and is subject to measurement error. Tenth, 44 newborns referred out or still admitted had no verifiable final outcome and were excluded, which would bias the mortality estimate if their outcomes differed systematically. Eleventh, the continuing-care components of the PLQS remain vulnerable to survivorship and opportunity bias, which is precisely why the early-care score was prespecified as primary and why the continuing-care result is not interpreted causally. Twelfth, the PLQS is study-defined and has not undergone external psychometric validation. Thirteenth, this is an observational analysis and the associations reported do not establish causation in either direction.

## Conclusion

Among preterm and low-birth-weight newborns admitted to eight facilities in Western Kenya, documented early patient-level quality of care was high, with a median early-care score of 93.5%, but was not associated with in-hospital mortality after adjustment for maturity, birth location, facility and condition at admission, and this null result was robust across nine sensitivity and bias-control analyses. Mortality was instead dominated by gestational age and by condition at presentation, particularly respiratory distress and hypothermia at admission. Specific indicators nonetheless require strengthening: sustained kangaroo mother care, individual thermal support rather than shared equipment, appropriate CPAP management and vitamin K administration were the weakest processes in these records. The marked difference between the null early-care result and the apparently protective association obtained when survival-dependent items were included in the same models indicates that composite quality scores spanning an entire admission can generate substantial spurious protective associations, and that quality-of-care research in this field should prespecify exposures that could reasonably have been delivered before the outcome occurred. These findings describe eight purposively assessed facilities and should not be generalised to newborn care in Kenya as a whole.

## Author contributions

MSS: Conceptualization, Methodology, Formal analysis, Data curation, Visualization, Writing – original draft. LNN: Conceptualization, Supervision, Writing – review & editing. CMS: Conceptualization, Funding acquisition, Project administration, Investigation, Writing – review & editing. TCK: Methodology, Validation, Writing – review & editing. PLW: Investigation, Data curation, Writing – review & editing. All authors read and approved the final manuscript.

## Data availability

The de-identified minimal dataset underlying the findings, with a data dictionary, the PLQS indicator definitions and a script that reproduces the main results, is openly available in Zenodo (https://doi.org/10.5281/zenodo.24297833) under a CC BY 4.0 licence. Names were not abstracted, and facility file numbers, all dates, facility names and free text were removed before deposition.

## Acknowledgments

The authors acknowledge the County Government of Bungoma Department of Health, the management and staff of the participating health facilities, the health records and information officers who supported record retrieval, and the research assistants who conducted the abstraction. Claude (Anthropic), a large language model, was used to assist with language editing and manuscript checks against journal requirements. The authors designed the study, collected and analysed the data, interpreted the findings, and reviewed, revised and take full responsibility for all content.

## Supporting information

**S1 Table. Missingness by analytical variable in the quality-of-care cohort.**

*(DOCX)*

**S2 Table. Facility-specific patient-level quality score summaries.**

*(DOCX)*

**S3 Table. Full model output for the primary and all sensitivity analysis populations, including the alternative exposure definitions.**

*(DOCX)*

**S4 Table. Interaction analyses with stratum-specific estimates.**

*(DOCX)*

**S5 Table. Full model output for the secondary outcome analyses.**

*(DOCX)*

**S6 Table. Model diagnostics.**

*(DOCX)*

**S1 Checklist. Completed STROBE checklist for cohort and cross-sectional studies.**

*(DOCX)*

