## Supplementary material for "Early patient-level quality of care and in-hospital outcomes among preterm and low-birth-weight newborns in Western Kenya: a multicentre retrospective record-review study": Online supplemental material

#### S1 Table. Missingness by analytical variable in the quality-of-care cohort

| Variable | Observed n | Missing n | Missing % |
|---|---|---|---|
| Gestational age (weeks) | 265 | 3 | 1.1 |
| Birth weight (g) | 267 | 1 | 0.4 |
| 5-minute Apgar | 231 | 37 | 13.8 |
| Multiple pregnancy | 255 | 13 | 4.9 |
| Mode of delivery | 268 | 0 | 0.0 |
| Maternal age | 213 | 55 | 20.5 |
| Antenatal visits | 193 | 75 | 28.0 |
| Hypothermia at admission | 253 | 15 | 5.6 |
| Hypoglycaemia at admission | 244 | 24 | 9.0 |
| Respiratory distress at admission | 260 | 8 | 3.0 |
| Suspected sepsis at admission | 260 | 8 | 3.0 |
| Disposition | 268 | 0 | 0.0 |
| Length of stay (days) | 247 | 21 | 7.8 |
| Discharge weight (g) | 224 | 44 | 16.4 |
| Feeding at discharge | 209 | 59 | 22.0 |

*Denominator is the quality-of-care cohort (n=268). Counts refer to values not recorded in the source records. Values not recorded for 5-minute Apgar score, multiple pregnancy, maternal age, antenatal visits and admission conditions were completed once during the independent data audit for use as covariates; gestational age was multiply imputed in the mortality models. Outcome variables and PLQS item scores were never statistically completed.*

Shisanya MS, et al. Early patient-level quality of care and in-hospital outcomes among preterm and low-birth-weight newborns in Western Kenya.

#### S2 Table. Facility-specific patient-level quality score summaries

| Facility | Newborns n | Early-care PLQS, median (IQR) | Continuing-care PLQS, median | Overall PLQS, median |
|---|---|---|---|---|
| Bumula Sub-County Hospital | 3 | 91.2 (88.2–92.6) | 89.3 | 87.1 |
| Bungoma County Referral Hospital | 80 | 86.7 (81.2–88.5) | 61.1 | 74.6 |
| Cheptais Sub-County Hospital | 22 | 73.0 (71.4–79.2) | 100.0 | 85.7 |
| Kabula Health Centre | 21 | 70.6 (61.8–84.4) | 83.3 | 74.2 |
| Kimilili Sub-County Hospital | 53 | 100.0 (100.0–100.0) | 100.0 | 100.0 |
| Naitiri Sub-County Hospital | 6 | 60.7 (60.2–65.2) | 65.7 | 64.3 |
| Sirisia Sub-County Hospital | 2 | 93.2 (92.8–93.7) | 97.9 | 95.4 |
| Webuye County Hospital | 81 | 100.0 (96.7–100.0) | 100.0 | 98.1 |

*PLQS, patient-level quality score; IQR, interquartile range. Facility case mix differs substantially and these summaries should not be read as a comparison of facility performance.*

Shisanya MS, et al. Early patient-level quality of care and in-hospital outcomes among preterm and low-birth-weight newborns in Western Kenya.

### S3 Table. Full model output for the primary and all sensitivity analysis populations

| Analysis population | n | Deaths | PLQS exposure, per 10 points | Gestational age, per week | Multiple pregnancy | Outborn | Referral hospital with newborn unit | 5-minute Apgar, per point | Hypothermia at admission |
|---|---|---|---|---|---|---|---|---|---|
| Primary (Model 3) | 221 | 56 | 1.02 (0.81–1.27) | 0.83 (0.77–0.89) | 1.04 (0.74–1.48) | 0.89 (0.53–1.47) | 1.93 (0.97–3.84) | 0.91 (0.81–1.03) | 1.77 (1.07–2.93) |
| Complete-case, Model 3 covariates | 221 | 56 | 1.02 (0.81–1.27) | 0.83 (0.77–0.89) | 1.04 (0.74–1.48) | 0.89 (0.53–1.47) | 1.93 (0.97–3.84) | 0.91 (0.81–1.03) | 1.77 (1.07–2.93) |
| Birth-weight category replacing gestational age | 223 | 57 | 0.94 (0.75–1.17) | — | 0.88 (0.63–1.23) | 0.82 (0.53–1.27) | 1.60 (0.80–3.20) | 0.93 (0.83–1.03) | 1.67 (1.03–2.69) |
| 24-hour survivor cohort | 215 | 50 | 1.03 (0.80–1.32) | 0.83 (0.77–0.90) | 1.02 (0.70–1.49) | 0.86 (0.49–1.52) | 2.13 (1.02–4.43) | 0.90 (0.78–1.04) | 1.81 (1.04–3.15) |
| At least 3 of 4 early-care domains valid | 221 | 56 | 1.02 (0.81–1.27) | 0.83 (0.77–0.89) | 1.04 (0.74–1.48) | 0.89 (0.53–1.47) | 1.93 (0.97–3.84) | 0.91 (0.81–1.03) | 1.77 (1.07–2.93) |
| Inborn newborns only | 163 | 37 | 1.09 (0.87–1.37) | 0.82 (0.76–0.89) | 0.88 (0.58–1.32) | — | 2.43 (1.06–5.60) | 0.73 (0.60–0.90) | 1.10 (0.60–2.05) |
| Outborn newborns only | 58 | 19 | 0.65 (0.38–1.12) | 0.85 (0.76–0.96) | 0.89 (0.36–2.21) | — | 0.76 (0.18–3.31) | 1.13 (0.92–1.38) | 3.05 (1.38–6.74) |
| Overall PLQS as exposure | 221 | 56 | 0.77 (0.66–0.90) | 0.86 (0.80–0.92) | 0.94 (0.64–1.37) | 0.87 (0.53–1.41) | 1.63 (0.82–3.24) | 0.88 (0.77–1.01) | 1.94 (1.18–3.19) |
| Continuing-care PLQS as exposure | 221 | 56 | 0.81 (0.74–0.89) | 0.88 (0.82–0.94) | 0.85 (0.57–1.27) | 0.91 (0.57–1.44) | 1.44 (0.76–2.74) | 0.87 (0.77–0.99) | 1.92 (1.19–3.10) |

*All values are adjusted risk ratios with 95% confidence intervals from modified Poisson regression with robust variance. A dash indicates the term was not present in that specification: birth location is excluded by design in the inborn-only and outborn-only populations, and gestational age is replaced by birth-weight category in that row. The final two rows substitute alternative exposure definitions into the identical specification in the same population and are the analyses demonstrating opportunity-to-receive-care bias. The main text reports one estimate per population; this table gives the full covariate output.*

Shisanya MS, et al. Early patient-level quality of care and in-hospital outcomes among preterm and low-birth-weight newborns in Western Kenya.

#### S4 Table. Interaction analyses with stratum-specific estimates

| Interaction | Newborns | Deaths | Ratio of risk ratios (95% CI) | p value | Stratum | Stratum n | Stratum deaths | Stratum adjuste ratio (95% CI) |
|---|---|---|---|---|---|---|---|---|
| Early-care PLQS × early preterm (<32 weeks) | 221 | 56 | 1.09 (0.66–1.80) | 0.735 | 32 weeks or more | 149 | 15 | 1.01 (0.60–1.70) |
| | | | | | Below 32 weeks | 72 | 41 | 1.05 (0.84–1.32) |
| Early-care PLQS × referred-in status | 221 | 56 | 1.00 (0.56–1.79) | 0.998 | Not referred in | 171 | 38 | 1.07 (0.76–1.51) |
| | | | | | Referred in | 50 | 18 | 0.73 (0.41–1.31) |

*CI, confidence interval; PLQS, patient-level quality score. Risk ratios are per 10-percentage-point increase in the early-care PLQS, estimated by modified Poisson regression with robust variance and adjusted for gestational age, multiple pregnancy, birth location, facility, 5-minute Apgar score, hypothermia at admission and suspected sepsis at admission. Birth location was omitted from the referral-status models. Both interaction analyses are underpowered and are reported as inconclusive*

Shisanya MS, et al. Early patient-level quality of care and in-hospital outcomes among preterm and low-birth-weight newborns in Western Kenya.

#### S5 Table. Full model output for the secondary outcome analyses

| Outcome and covariate | Estimate | 95% confidence interval | p value | Effect measure |
|---|---|---|---|---|
| **Any complication during admission (n = 265)** | | | | |
| Early-care PLQS, per 10 points | 1.01 | (0.91–1.11) | 0.916 | Adjusted risk ratio |
| Gestational age, per week | 0.96 | (0.94–0.98) | <0.001 | Adjusted risk ratio |
| Multiple pregnancy | 1.01 | (0.85–1.20) | 0.951 | Adjusted risk ratio |
| Outborn | 1.45 | (1.21–1.74) | <0.001 | Adjusted risk ratio |
| Referral hospital with newborn unit | 3.76 | (2.45–5.77) | <0.001 | Adjusted risk ratio |
| 5-minute Apgar, per point | 0.95 | (0.90–0.99) | 0.020 | Adjusted risk ratio |
| **Sepsis during admission (n = 265)** | | | | |
| Early-care PLQS, per 10 points | 0.59 | (0.49–0.71) | <0.001 | Adjusted risk ratio |
| Gestational age, per week | 1.03 | (0.98–1.09) | 0.284 | Adjusted risk ratio |
| Multiple pregnancy | 1.00 | (0.70–1.41) | 0.987 | Adjusted risk ratio |
| Outborn | 1.65 | (1.10–2.46) | 0.015 | Adjusted risk ratio |
| Referral hospital with newborn unit | 19.11 | (4.88–74.82) | <0.001 | Adjusted risk ratio |
| 5-minute Apgar, per point | 1.01 | (0.94–1.08) | 0.836 | Adjusted risk ratio |
| **Referral out of the facility (n = 265)** | | | | |
| Early-care PLQS, per 10 points | 0.68 | (0.56–0.82) | <0.001 | Adjusted risk ratio |
| Gestational age, per week | 0.91 | (0.81–1.02) | 0.113 | Adjusted risk ratio |
| Multiple pregnancy | 0.71 | (0.29–1.74) | 0.459 | Adjusted risk ratio |
| Outborn | 1.62 | (0.76–3.47) | 0.210 | Adjusted risk ratio |
| Referral hospital with newborn unit | 0.00 | (0.00–0.00) | <0.001 | Adjusted risk ratio |
| 5-minute Apgar, per point | 0.79 | (0.64–0.99) | 0.040 | Adjusted risk ratio |
| **Human-milk feeding at discharge (n = 160)** | | | | |
| Early-care PLQS, per 10 points | 0.96 | (0.93–0.98) | 0.003 | Adjusted risk ratio |
| Gestational age, per week | 1.02 | (1.00–1.05) | 0.059 | Adjusted risk ratio |
| Multiple pregnancy | 0.94 | (0.84–1.04) | 0.223 | Adjusted risk ratio |
| Outborn | 0.97 | (0.85–1.11) | 0.646 | Adjusted risk ratio |
| Referral hospital with newborn unit | 1.16 | (1.05–1.30) | 0.005 | Adjusted risk ratio |
| 5-minute Apgar, per point | 1.00 | (0.97–1.03) | 0.932 | Adjusted risk ratio |
| **Average daily weight gain, g/day (n = 160)** | | | | |
| Early-care PLQS, per 10 points | -1.6 | (-4.8 to 1.5) | 0.304 | Adjusted mean difference |
| Gestational age, per week | -0.4 | (-1.7 to 1.0) | 0.582 | Adjusted mean difference |
| Multiple pregnancy | -6.2 | (-13.5 to 1.2) | 0.102 | Adjusted mean difference |
| Outborn | 13.6 | (2.0 to 25.2) | 0.022 | Adjusted mean difference |
| Referral hospital with newborn unit | -19.9 | (-28.0 to -11.8) | <0.001 | Adjusted mean difference |
| 5-minute Apgar, per point | -0.1 | (-2.3 to 2.1) | 0.927 | Adjusted mean difference |
| **Weight change, g (n = 160)** | | | | |
| Early-care PLQS, per 10 points | -17.5 | (-40.8 to 5.8) | 0.141 | Adjusted mean difference |
| Gestational age, per week | -34.8 | (-52.5 to -17.1) | <0.001 | Adjusted mean difference |

| Outcome and covariate | Estimate | 95% confidence interval | p value | Effect measure |
|---|---|---|---|---|
| Multiple pregnancy | 14.5 | (-51.7 to 80.8) | 0.667 | Adjusted mean difference |
| Outborn | 139.6 | (40.4 to 238.8) | 0.006 | Adjusted mean difference |
| Referral hospital with newborn unit | -50.2 | (-115.8 to 15.3) | 0.133 | Adjusted mean difference |
| 5-minute Apgar, per point | -20.9 | (-43.1 to 1.2) | 0.064 | Adjusted mean difference |

*All models adjust for the covariates shown. Binary outcomes were analysed by modified Poisson regression; weight outcomes by linear regression with robust standard errors. Table 5 in the main text reports only the PLQS exposure estimate for each outcome; this table gives full covariate output. All secondary analyses are exploratory.*

Shisanya MS, et al. Early patient-level quality of care and in-hospital outcomes among preterm and low-birth-weight newborns in Western Kenya.

#### S6 Table. Model diagnostics

| Diagnostic | Assessment | Action taken |
|---|---|---|
| Exposure distribution | Early-care PLQS strongly left-skewed; median 93.5%, interquartile range 83.3–100; 37.9% of the mortality cohort scored exactly 100 | Modelled continuously per 10 points and additionally in four categories defined by the observed distribution, with the ceiling category separated |
| Nonlinearity | Quadratic term risk ratio 0.91 (95% CI 0.66–1.25), p=0.555; restricted cubic spline term with knots at the 10th, 50th and 90th centiles, p=0.301 | No evidence of departure from linearity; spline model not adopted and no spline figure fitted |
| Collinearity: gestational age and birth weight | Strongly correlated markers of maturity | Gestational age used in the primary adjustment set; birth-weight category substituted in a prespecified sensitivity model; the two never entered together |
| Sparse strata: facility | Deaths concentrated in three of eight facilities; the remaining five contributed 34 newborns and no deaths | Individual facility fixed effects produced complete separation; facility modelled as a binary term contrasting the two referral hospitals with newborn units against all other facilities |
| Separation: respiratory distress at admission | Documented in 121 of 224 newborns; 1 of 103 newborns without it died, against 56 of 121 with it | Excluded from the primary Model 3; crude and forced-entry adjusted estimates reported separately with explicit note of imprecision |
| Events per parameter | 57 deaths; Model 2 five parameters (11.4 events per parameter); Model 3 eight parameters (7.1 events per parameter) | Model 3 reduced from the originally planned specification by the prespecified conceptual hierarchy rather than by any p-value-driven procedure |
| Model convergence | All modified Poisson models converged within 100 iterations | No convergence failures; a random-intercept facility model was not fitted, since events in five of eight facilities were zero |
| Overdispersion: length of stay | Mean 15.8 days, variance 977.3 among discharged survivors | Poisson regression with robust variance used to estimate an adjusted ratio of means rather than ordinary linear regression |
| Influential observations: weight outcomes | Implausible average daily weight gain values identified in the raw distribution | Weight analyses restricted to discharged survivors with valid paired weights and biologically plausible daily change |
| Missing covariate data | Gestational age missing in 3 of 224 (1.3%) in the mortality cohort; no other model covariate incomplete | Twenty imputations by chained equations with Rubin's rules pooling; complete-case analysis reported alongside and estimates compared directly |

*CI, confidence interval; PLQS, patient-level quality score*

Shisanya MS, et al. Early patient-level quality of care and in-hospital outcomes among preterm and low-birth-weight newborns in Western Kenya.

### S1 Checklist. Completed STROBE checklist for cohort and cross-sectional studies

| Item | Requirement (abbreviated) | Location in the manuscript |
|---|---|---|
| STROBE 1 | Study design indicated in title or abstract; informative summary | Title; Abstract |
| STROBE 2 | Scientific background and rationale | Introduction |
| STROBE 3 | Specific objectives | Introduction, final paragraph |
| STROBE 4 | Key elements of study design presented early | Methods, study design |
| STROBE 5 | Setting, locations and relevant dates | Methods, setting and data sources |
| STROBE 6 | Eligibility criteria, sources and methods of selection | Methods, study population |
| STROBE 7 | Variables defined, including outcomes, exposures and confounders | Methods, variable and outcome definitions |
| STROBE 8 | Sources of data and methods of measurement | Methods, data sources |
| STROBE 9 | Efforts to address potential sources of bias | Methods, bias handling; Discussion, limitations |
| STROBE 10 | Study size explained | Results, cohort derivation; flow figure |
| STROBE 11 | Quantitative variables and groupings handled | Methods, variable definitions |
| STROBE 12 | Statistical methods, subgroups, missing data, sensitivity analyses | Methods, statistical analysis; missing data; sensitivity analyses |
| STROBE 13 | Participant flow and numbers at each stage | Results, cohort derivation; flow figure |
| STROBE 14 | Descriptive data and missingness | Characteristics table; missingness supplementary table |
| STROBE 15 | Outcome data reported | Main results tables |
| STROBE 16 | Unadjusted and adjusted estimates with precision | Regression tables; supplementary full model output |
| STROBE 17 | Other analyses, including subgroups and sensitivity analyses | Results, sensitivity analyses; supplementary tables |
| STROBE 18 | Key results summarised with reference to objectives | Discussion, principal findings |
| STROBE 19 | Limitations discussed, including bias and imprecision | Discussion, limitations |
| STROBE 20 | Cautious overall interpretation | Discussion throughout; Conclusion |
| STROBE 21 | Generalisability | Discussion, limitations; Conclusion, final sentence |
| STROBE 22 | Funding source and role | Funding statement |

Shisanya MS, et al. Early patient-level quality of care and in-hospital outcomes among preterm and low-birth-weight newborns in Western Kenya.
